# Pharmacogenomic Diversity in Psychiatry: Challenges and Opportunities in Africa

**DOI:** 10.1101/2024.01.16.24301341

**Authors:** Muktar B. Ahmed, Anwar Mulugeta, Niran Okewole, Klaus Oliver Schubert, Scott R. Clark, Conrad O. Iyegbe, Azmeraw T. Amare

## Abstract

Pharmacogenomic studies on psychiatric drugs have slowly identified genetic variations that influence drug metabolism and treatment effectiveness in patients with mental illness. However, most of these studies have predominantly centered on people of European descent, leaving a substantial knowledge gap on the clinical implications of current pharmacogenomic evidence in multi-ancestry populations such as Africans. Thus, whether pharmacogenomic (PGx) genetic testing implemented in European populations would be valid for a population of African origin is unknown. The objective of this review was to appraise previous psychiatric pharmacogenomic studies in Africa and highlight challenges and opportunities to initiate PGx testing in the region. A systematic literature search was conducted on PubMed, Scopus, and Web of Science to identify studies published in the English language up to January 26, 2024. The primary outcomes were treatment response, remission, side effects, and drug metabolism in African psychiatric patients.

The review included 42 pharmacogenomic studies that explored the genetic profiles of psychiatric patients in Africa. Despite the limited number of studies, our review found strong evidence of pharmacogenomic diversity within the African populations, emphasizing the importance of pharmacogenomic research in this population. A high degree of variability and differences in the frequencies of cytochrome P450 (CYPs) genotypes have been reported within the African population. It is estimated that 28% of North Africans and Ethiopians are ultrarapid metabolizers of several medications, mainly attributed to the increased activity of the *CYP2D6* enzyme. This prevalence is significantly higher than that among Caucasians (10%), or Hispanics, Chinese, or Japanese populations (1%). Due to the defective *CYP2C19*2* allele (at a frequency of 14%) and *CYP2C19*3* allele (2% frequency), 5.2% of Ethiopians were identified as poor metabolizers of S-mephenytoin, a probe substrate used to assess the activity of the cytochrome P450 enzyme. In Tunisian patients with schizophrenia, genotyping the *CYP1A2* gene and using therapeutic drug monitoring (TDM) improved the effectiveness and safety of clozapine. Among South African patients with schizophrenia, antipsychotic treatment response was associated with two gene variants (rs13025959 in the *MYO7B* gene with the ‘C’ allele and rs10380 in the *MTRR* gene with the ‘T’ allele).

Overall, the review has identified evidence of pharmacogenomic diversity in African populations and recommended expanding pharmacogenomic studies while introducing PGx testing in this population. For the few characterized genes, Africans showed qualitative and quantitative differences in the profile of pharmacogenetic variants when compared to other ethnic groups. Limited research funding, inadequate infrastructure, and a shortage of skilled human resources might be a challenge, but by building upon local successes and through collaborations with international partners, it is possible to establish pharmacogenomic biobanks and leverage global genetic resources to initiate personalized treatment approaches in Africa.

## Introduction

Psychiatric disorders continue to be a major global concern, ranking among the top ten leading causes of disease burden worldwide [1]. In 2019, mental disorders accounted for 125.3 million disability-adjusted life years (DALYs) globally, representing 4.9% of the global burden of disease [1]. The prevalence of mental health disorders is a significant challenge in Africa, given the region’s socioeconomic and political adversities [2]. According to a scoping review of 36 studies from 12 African countries, lifetime prevalence rates of mood disorders range from 3.3% to 9.8%, anxiety disorders range from 5.7% to 15.8%, substance use disorders range from 3.0% to 13.3%, and psychotic disorders range from 1.0% to 4.4% [3].

To address the significant prevalence of mental health disorders, access to pharmacotherapy is widely promoted, and various initiatives have been implemented to enhance the availability of mental healthcare services and medications throughout Africa [4]. However, the availability of pharmacotherapy does not guarantee the attainment of the most favorable health outcomes, as it necessitates prescribing the right medication for the right patient. For example, in studies of largely Caucasian populations, 30-35% of patients with mental illness who receive pharmacological therapy fail to respond to first-line treatment [5], partly attributed to the impact of genetic factors on drug metabolism and treatment effectiveness. Genetic factors contribute 30-40% of the interindividual variability in response to mood stabilizers and antipsychotics [6, 7] and specifically common genetic variants account for up to 10% of the variance in the plasma concentrations of clozapine and its metabolites [8]. Furthermore, treatment effectiveness in patients with psychiatric disorders is influenced by a complex interplay between genetic and environmental factors. For instance, personality traits [9], and stressful life events reported during or immediately before treatment [10] predict poor response.

To date, pharmacogenomic studies have successfully identified genetic variations involved in the metabolism of various psychotropic drugs (e.g., *CYP2D6* and *CYP2C19*) [11] and drug transporters (e.g., *5-HTTLPR),* establishing their association with treatment outcomes in psychiatry, including response [12], remission [13], resistance [14] and adverse drug reactions [15]. Genetic polymorphisms located within or near pharmacologically relevant candidate genes and combined scores from these variants [16–20] have shown association with individuals’ reactions to psychiatric medications [21]. In patients with bipolar disorder (BD), a high polygenic loading for schizophrenia (SCZ) or major depressive disorder (MDD) is significantly associated with poor response to lithium treatment [22–24], and incorporating clinical variables in conjunction with the polygenic scores has enhanced the ability to predict the response to lithium treatment in these patients [25–27]. Similarly, in patients with MDD, the polygenic scores for neuroticism and openness personality traits were associated with the likelihood of remission to selective serotonin reuptake inhibitors (SSRIs) treatment [28].

The large majority of these studies; however, are conducted in developed countries mainly using samples of European ancestry, posing a challenge to the potential transferability of existing pharmacogenomic (PGx) testing platforms to other settings, such as in Africa [29–32]. To gain a more profound perspective into the scope of psychiatric pharmacogenomics in African populations, we conducted a systematic review and assessed potential prospects and challenges that could advance psychiatric PGx testing within the African populace. We also explored current challenges and opportunities and highlighted how the global community could benefit from leveraging the pharmacogenomics diversity of Africans.

## Materials and Method

### Search strategy

Using PubMed, SCOPUS, and Web of Science databases, we conducted a comprehensive search of all pharmacogenomic studies published until 26/01/2024. In PubMed, we employed Medical Subject Headings (MeSH) and text word search strategies. For SCOPUS, we utilized Emtree (EMBASE vocabulary) terms along with text word searches. Our search strategy involved combining broad keywords such as “pharmacogenetics”, “pharmacogenomics”, and “psychiatric disorders” with specific search terms adapted to each database and limited to research in African settings. Search terms related to pharmacogenomics included “pharmacogenetics-based dosing” and “pharmacogenomic testing,” while terms related to psychiatric disorders encompassed “mental illness,” “psychological disorders,” and “behavioral disorders.” To ensure comprehensiveness, we incorporated synonymous and related terms for each keyword, combining them using Boolean operators “AND” and “OR”. Our search strategy included not only electronic database searches but also hand-searches of the references included in the study. Finally, using an excel sheet, we extracted relevant information from each study including candidate genes, treatment outcomes, the type of psychiatric disorders, and the study population. The findings of this systematic review were reported in accordance with the reporting guidelines of Preferred Reporting Items for Systematic Review and Meta-Analyses (PRISMA) [33].

### Selection criteria and data extraction

The review process including article selection and evaluation was conducted independently by two reviewers (MA and AA). Articles conducted in African populations and focusing on pharmacogenomics or pharmacogenetics of psychiatric disorders were screened using the predefined eligibility criteria. We excluded studies that did not involve human subjects, were not published in English, or for which the full text was not available. Disagreements between MA and AA were resolved by discussion. All retrieved records were exported into Endnote library (version 20) [34] for further analysis.

We utilized a customized data extraction template to collect relevant information, including authors, publication year, sample size and ancestry characteristics, type of psychiatric condition and medications used, pharmacogenomic outcomes, and reported genes/alleles. Further details regarding the search strategy, inclusion and exclusion criteria, and results of the literature search are provided in the Supplementary file.

*Insert supplementary file here*

## Results

Our initial literature search identified 1767 articles. After removing duplicates, 1090 remained and were screened based on title and abstract. Subsequently, 42 studies (35 on pharmacogenetics and 7 on pharmacogenomics) were included in the final synthesis. Figure 1 shows the flow diagram of the literature review process.

**Figure 1.**
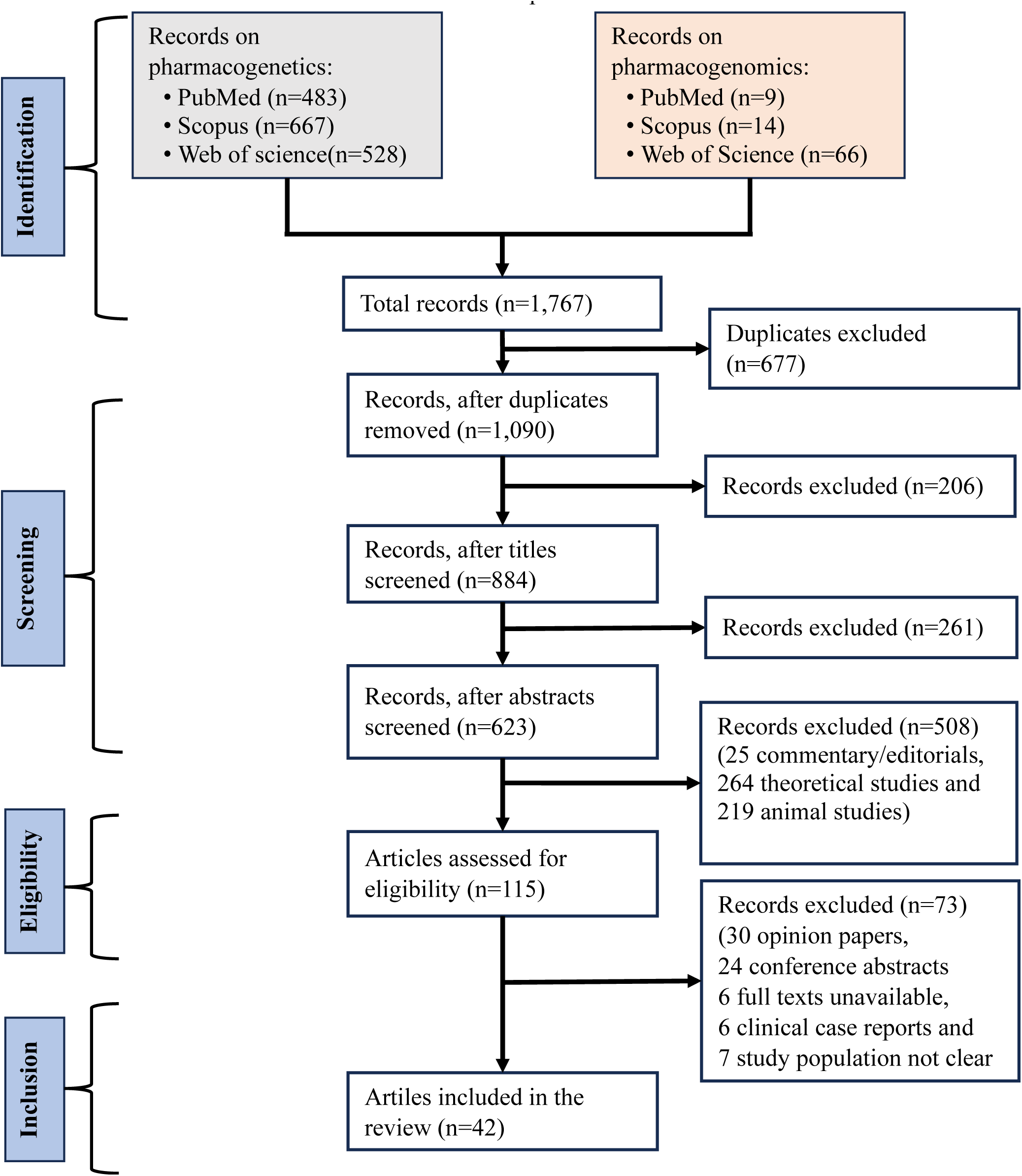
A flow diagram illustrating the stepwise screening of articles on the pharmacogenomic of psychiatric disorders in the African population. Our review found a considerable variation in the distribution of psychiatric pharmacogenomic studies across Africa. Countries such as South Africa, Zimbabwe, Uganda, Tunisia, Ghana, and Algeria were more strongly represented than others. Of all the studies reviewed, the largest proportion of the studies (23%) were conducted in southern African regions, followed by the eastern (20%) and Northern African regions (12%). The West African region was represented by approximately 9% of the studies, while the Central African region had the smallest proportion, accounting for less than 1% of the total. It has been reported that 35% of the studies involved individuals from multiple African regions (Figure 2).

**Figure 2.**
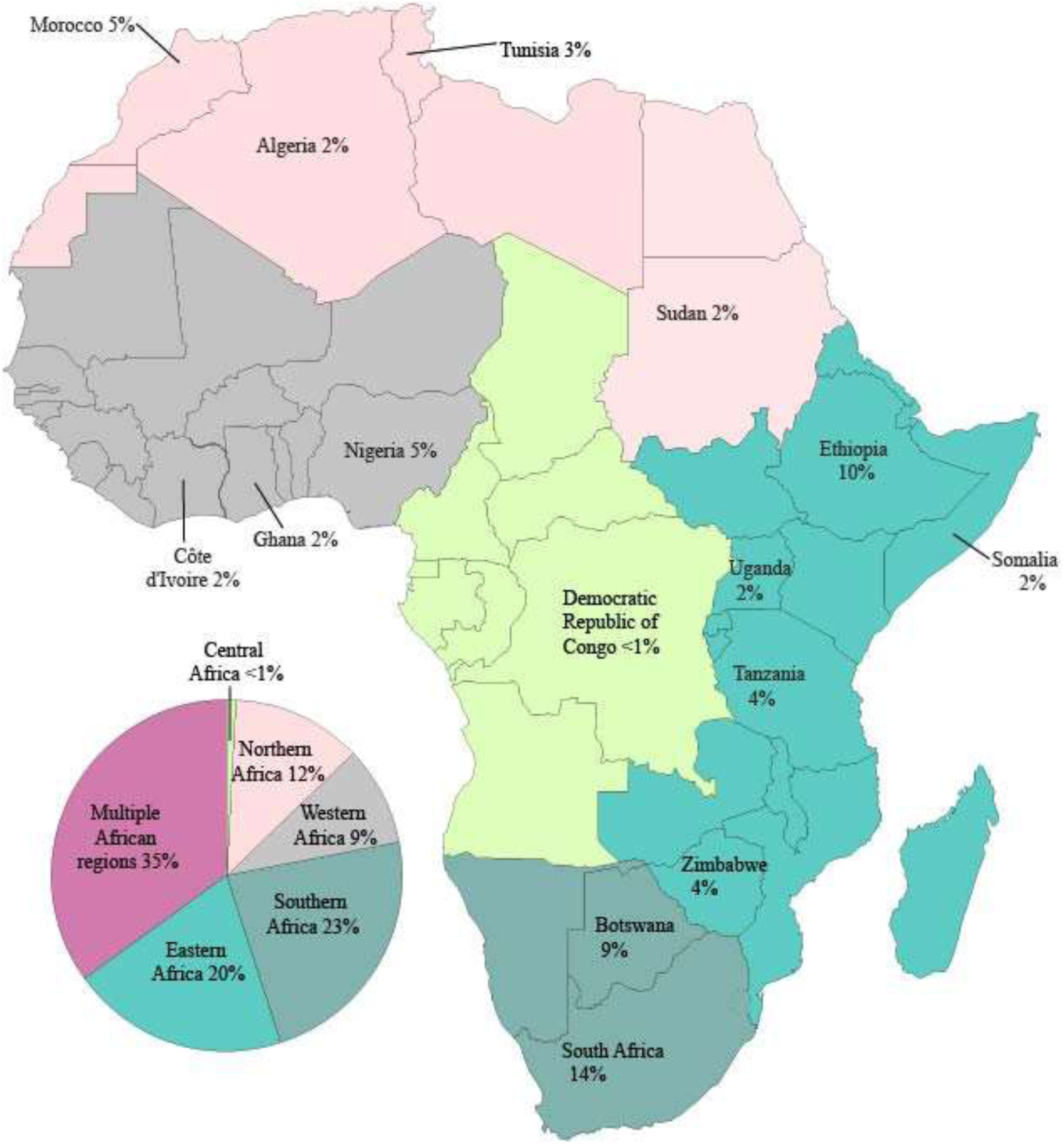
Distribution of pharmacogenomic and pharmacogenetic studies in patients with psychiatric disorders across African regions

### Psychiatric Pharmacogenomics in African Populations

Pharmacogenomic research in African populations has predominantly centered on Cytochrome P450 (CYPs), a group of enzymes that play a critical role in drug metabolism [35], including psychiatric medications (see Table 1). By primarly focusing on patient with SCZ or MDD, these studies revealed substantial pharmacogenomic diversity in the core ADME (Absorption, Distribution, Metabolism, and Excretion) and extra ADME genes[36]. This diversity was observed both within Africans and across individuals of African and European ancestry, emphasizing the need for tailored PGx testing programs that accommodate population-specific genetic variations [36].

**Table 1.** Psychiatric pharmacogenomics among individuals of African ancestry.

| Author<br>(Year) | Country (N) | Drug | Treatment<br>outcome<br>(Phenotype) | Gene | Psychiatric<br>disorder | Summary of finding |
| --- | --- | --- | --- | --- | --- | --- |
| **Benmessaoud, D. et al., (2008)**[47] | Algeria (88) | Antipsychotics (mainly Haloperidol) for 4 weeks | Treatment-refractory schizophrenia (TRS) | G allele of the -1438A/G 5-*HTR2A* | SCZ | The -1438A/G polymorphism 5-*HT2A* receptor gene showed association with a favourable response to typical antipsychotics. The ‘G’ allele identified individuals with SCZ who have a higher likelihood of responding to antipsychotics. |
| **Campbell, D. B. et al., (2008)**[38] | Clinical Antipsychotic Trials of Intervention Effectiveness (CATIE). Out of 678 subjects 198 were African only | Perphenazine and Quetiapine | Antipsychotic treatment efficacy, assessed using Positive and Negative Symptoms Scale (PANSS) | *RGS4* (rs2661319, rs2842030) for baseline PANSS total score. rs951439 ‘C’ allele for treatment response in African ancestry | SCZ | *RGS4* genotypes predicted treatment response. For individuals of African ancestry with rs951439 ‘C’ allele homozygosity, perphenazine was more effective than quetiapine (p=0.010) or ziprasidone (p=0.002). |
| **Drögemöller et al., (2015)**[40] | South Africa (190) | Flupentixol decanoate | PANSS scale | rs13025959 in *MYO7B* (E1647D) and rs10380 in *MTRR* (H622Y) | First episode SCZ | rs13025959 in *MYO7B* gene was linked to a reduced progress in overall psychopathological symptoms following antipsychotic treatment. rs13080 in *MTRR* was associated with an improvement in positive symptoms after antipsychotic treatment. |
| **Ammar, H. et al., (2021)[46]** | Tunisia (51) | Clozapine | Therapeutic drug monitoring (Variability in mean dose adjustment of clozapine). | *CYP1A2\*1F* (rs762551; -163C>A), *CYP1A2\*1C* (rs2069514; -3860 G>A) and *CYP2C19\*2* (rs4244285; 681G>A) | SCZ | *CYP1A21\*F* and *CYP1A2* -163C>A variants were linked to variation in Clozapine exposure among Tunisian patients with SCZ. *CYP1A21\*F* alone explained 24% of the variability in Clozapine C0/D. The authors recommended genotyping *CYP1A2* and using therapeutic drug monitoring (TDM) to improve Clozapine's effectiveness and safety in treating SCZ patients. |
| **Drögemöller et al., (2014)[43]** | Mixed ancestry of first episode SCZ (n=103) and Xhosa (n=222) population in South Africa | Antipsychotic | Treatment response | rs11368509 | SCZ | A frameshift variant, rs11368509 in *UPP2* found to be associated with improved treatment response to antipsychotics in mixed ancestry (p=0.057) and South African Xhosa populations (p=0.016) |
| **Ovenden, E. S. et al., (2017)[42]** | South African (103) | Flupentixol decanoate | Percentage change in PANSS | *MANBA*, *COL9A2* and *NFKB1* | SCZ | Noncoding variants on *MANBA*, *COL9A2*, and *NFKB1* were examined for their treatment response in antipsychotic response. Three SNPs (rs230493, rs3774959, and rs230504) were associated with poor treatment response in SCZ. |
| **O'Connell, K. S., et al., (2019)[41]** | South African (20) | Flupentixol decanoate | Differential expression of regulatory genes to identify candidate genes for | (*DNMT3A* rs2304429, *HDAC5* rs11079983, and *HDAC9* rs1178119 and rs11764843 | SCZ | Four variants within differentially expressed genes (*DNMT3A* rs2304429, *HDAC5* rs11079983, and *HDAC9* rs1178119 and rs11764843) were significantly associated with treatment response in SCZ. Two of these variants altered the expression of specific genes in |
|  |  |  | association with antipsychotic treatment response. |  |  | brain regions previously implicated in SCZ and treatment response, potentially aiding in the development of biomarkers and novel drug targets for antipsychotic treatment response. |
| **Antonio F Pardinas et al., (2023)**[37] | CLOZUK study (192) | Clozapine | Clozapine and norclozapine plasma concentrations | *for Clozapine*<br>[DNAJB3, MROH2A, UGT1A1, UGT1A3, UGT1A4, UGT1A5, UGT1A6, UGT1A7, UGT1A8, UGT1A9, UGT1A10, CYP1A1, CYP1A2];<br>*for Norclozapine*<br>[UGT2A3, UGT2B4, UGT2B7, UGT2B10, UGT2B11, UGT2B28, DNAJB3, MROH2A, UGT1A1, UGT1A3, UGT1A4, UGT1A5, UGT1A6, UGT1A7, UGT1A8, UGT1A9, UGT1A10, CYP1A1, CYP1A2] | SCZ | Individuals of sub-Saharan African ancestry showed lower levels of clozapine (beta -0.088) and norclozapine (beta -0.280) in their plasma compared to Europeans, indicating that they metabolize these drugs faster. |

### Pharmacogenomics in patients with Schizophrenia

Studies involving Africans diagnosed with SCZ have identified several genetic variations that play a role in pharmacotherapeutic outcomes. These include *CYP1A2* (CYP1A21* F, -163C>A), *RGS4* (rs2661319, rs2842030, rs951439), *UPP2* (rs11368509), *CNR1* (rs1049353), *MYO7B* (rs13025959), *MTRR* (rs10380), *DNMT3A* (rs2304429), *HDAC5* (rs11079983), *HDAC9* (rs11764843, rs1178119), *GRM3* (rs724226), *COMT* (rs165599), *DRD2* (rs1801028, Taq1A, Taq1B, rs1125394) and NFKB1 (rs230493, rs230504, and rs3774959). The CLOZUK study found that people with sub-Saharan African ancestry metabolized clozapine and norclozapine faster than people with European ancestry [37]. In individuals of African ancestry, the presence of genetic variants within *RGS4* gene (rs2661319 and rs2842030) predicted relative responsiveness to atypical antipsychotic treatment (perphenazine). Those who were homozygous for ‘C’ allele of rs951439 in *RGS4* gene had shown a more favourable response to perphenazine compared to quetiapine or ziprasidone [38].

In patients with first-episode of SCZ from Xhosa populations in South Africa, frameshift variant, rs11368509 in *UPP2* gene, was associated with improved anti-psychotic response [39]. In another study, rs13025959 in the *MYO7B* gene and rs10380 in the *MTRR* gene were associated with different aspects of treatment response. The ’C’ allele of rs13025959 (*MYO7B*) negatively affected improvement in general psychopathological symptoms after antipsychotic treatment, whereas the ’T’ allele of rs10380 (*MTRR*) was associated with favourable treatment outcome [40]. The *DNMT3A* gene variant rs2304429 ‘CC’ genotype was associated with an improved antipsychotic treatment response in South African SCZ patients compared to those who carry the ‘TC’ allele [41]. In the same study, *HDAC5* rs11079983 ‘TT’ genotype was associated with a poorer treatment response when compared to the ‘CC’ genotype [41]. Patients with the *HDAC9* rs11764843 ‘CA’ or *HDAC9* rs1178119 ‘GA’ genotype had a slower rate of reduction in negative symptom scores over time on the Positive and negative Syndrome Scale (PANSS) than patients with the ‘AA’ genotype [41]. A similar study in South African patients with SCZ examined the association of noncoding genetic variants within the *NFKB1* with treatment response. Of the three SNPs identified, rs230493 and rs230504 were associated with higher PANSS negative symptoms score, while rs3774959 was associated with higher PANSS negative and PANSS total scores [42]. These variants located in the 4q24 region, which characterised by XXXXX.

In African American patients, the synonymous polymorphism (rs1049353) in *CNR1* gene was associated with a higher weight gain following clozapine and olanzapine treatment. Allelic analysis of rs1049353 (‘CC’ vs ‘CT’ allele) demonstrated that patients who carried the ‘C’ allele gained more weight compared to those with ‘T’ allele [43]. Genetic variants in *GRM3, COMT,* and *DRD2* were associated with changes in PANSS scores in response to risperidone treatment (atypical antipsychotic). In those taking risperidone, the *GRM3* SNP-rs724226 was associated with change in the PANSS total score, while the *COMT* SNP-165599 was moderately associated with change in the PANSS negative score. In addition, there was a significant association between rs1801028, a SNP in *DRD2*, and PANSS negative symptoms. Moreover, the change in the PANSS negative total score increased with a higher number of ‘G’ alleles among African-American patients with ‘AA’, ‘AG’, and ‘GG’ genotypes [44]. *DRD2* variants, such as *Taq1A*, *Taq1B*, and rs1125394, have also been associated with an improved response to clozapine[45]. For example, significant differences in the frequencies of the *Taq1B* allele were observed between responders and non-responders to clozapine, with the “B2” allele being associated with a better response. There were also significant differences in allele frequencies at two other marker sites, with allele 1 of rs1125394 and allele A2 of Taq1A being more frequent among responders. However, no significant association was found between clozapine response and insertion/deletion (ins/Del) site -141C [45].

Genotyping the *CYP1A2* gene was found to be associated with improved effectiveness and safety of Clozapine in Tunisian patients with SCZ [46]. This study found a significant association between the *CYP1A21* F* polymorphism and Clozapine metabolism, accounting for 24% of the variability in Clozapine concentration-to-dose ratio. Moreover, the *CYP1A2 - 163C>A* variant plays a crucial role in influencing the plasma levels of Clozapine. This study suggested that integrating *CYP1A2* genotyping with therapeutic drug monitoring (TDM) of Clozapine might enhance both efficacy and safety, although further validation of these findings is warranted [46]. In Algerian patients with SCZ, those who carry the ’G’ allele for a gene that codes for the 5-HT2A receptor showed a more favourable response to conventional antipsychotics [47].

### Pharmacogenomics in patients with depression

In Africans with depression, studies revealed the influence of genetic variations such as *CYP2C19* (CYP2C19*2), *CYP2D6* (2509G>T in CYP2D6*2/*29 genotype) and *CRHBP* (rs10473984) on the metabolism and treatment effectiveness of antidepressants. For instance, a case report utilizing data from a sub-Saharan African patient with MDD identified the presence of the *CYP2C19*\*2 loss of function allele. This varation was observed following voriconazole treatment for *Aspergillus fumigatus* renal abscess and co-administration of haloperidol, a weak *CYP3A4* inhibitor [48]. In African Americans, it was discovered that the variant rs10473984 within the *CRHBP* locus was associated with both depressive symptoms reduction and remission in patients taking citalopram, particularly those with features of anxious depression. In this study, the rs10473984 ’T’ allele was associated with poorer treatment outcomes[49]. Another study reported a significant relationship between a genetically determined African ancestry or self-reported race and poor treatment response to citalopram [50]. A study on South African patients taking amitriptyline found that a genetic variant, *2509G>T* in *CYP2D6*2/*29* genotype resulted in an amino acid change (*K245N*) and amitriptyline treatment-related side effects [51].

### Pharmacokinetic genes within the African population

Table 2 summarises the pharmacokinetic and genetic profile studies conducted on African population. The findings reveal a notable genetic variations wthin candidate gene that impact the metabolism of psychiatric medications and their treatment outcomes in African patients. Studies conducted in Tanzania, Zimbabwe, Sudan, Somalia, Burundi, Ethiopia, and South Africa demonstrated significant differences in the frequency of various CYP family genes such as *CYP2D6*, *CYP2C19*, *CYP3A4*, *CYP2B6*, *CYP2*, and *CYP1A2* [29]. Many of the pharmacogenes studied in the African population belong to the highly polymorphic gene of *CYP2D6*, which is involved in the metabolism of up to 25% of commonly prescribed psychiatric medications [52]. A study focused on *CYP2D6* genetic variants related to tramadol treatment (a prodrug, metabolized in liver) in an African population revealed a significant difference in the maximum plasma concentrations of the active metabolite (+) R, R-O-desmethyltramadol between the ultra-metabolisers (UMs) and extensive metabolisers (EMs). UMs showed an increased pain threshold and tolerance compared to EMs and nearly 50% of the UM group experienced nausea compared to only 9% of the EM group [53]. For instance, in a study of Tanzanian psychiatric patients, healthy Tanzanian controls, and South African Venda populations, the *CYP2D6* genotype was found to predict poor metabolisers in only a small percentage of cases 2.3%, 1.9%, and 2.6%, respectively. Notably, in the Tanzanian population, the frequency of the low-activity *CYP2D6*17* allele was significantly higher among psychiatric patients (30%) compared to healthy individuals (20%). In healthy Tanzanian individuals, EMs showed a significantly higher sparteine metabolic ratio (SMR, a pharmacogenetic measure of *CYP2D6* activity) compared to white Danish EMs. The frequencies of the *CYP2D6*1* and *CYP2D6*4* alleles were 9.6% and 4.0%, in Tanzanians and white Danish samples respectively, and no *CYP2D6*3* alleles were found. The frequencies of the *CYP2C19*\*1 and *CYP2C19*2* alleles was 9.0%, 1.0%, respectively [54]. Moreover, a study identified a unique allele distribution and two rare novel alleles, *CYP2D6*\*73 and *CYP2D6*\*74 in South African participants [55]. With this pharmacogene, genotyping of black Zimbabwean population on *CYP2D6* showed that the frequency of three genes, *CYP2D6*A*, *CYP2D6*B*, and *CYP2D6*D*, was determined to be 0%, 1.8%, and 3.9%, respectively. None of the subjects carried the Xba I 44 kb haplotype, which is a known *CYP2D6* allele indicative of poor metabolisers in Caucasians [56]. The *CYP2D6*B* allele frequency was very low in the Zimbabwean population [57].

**Table 2.**
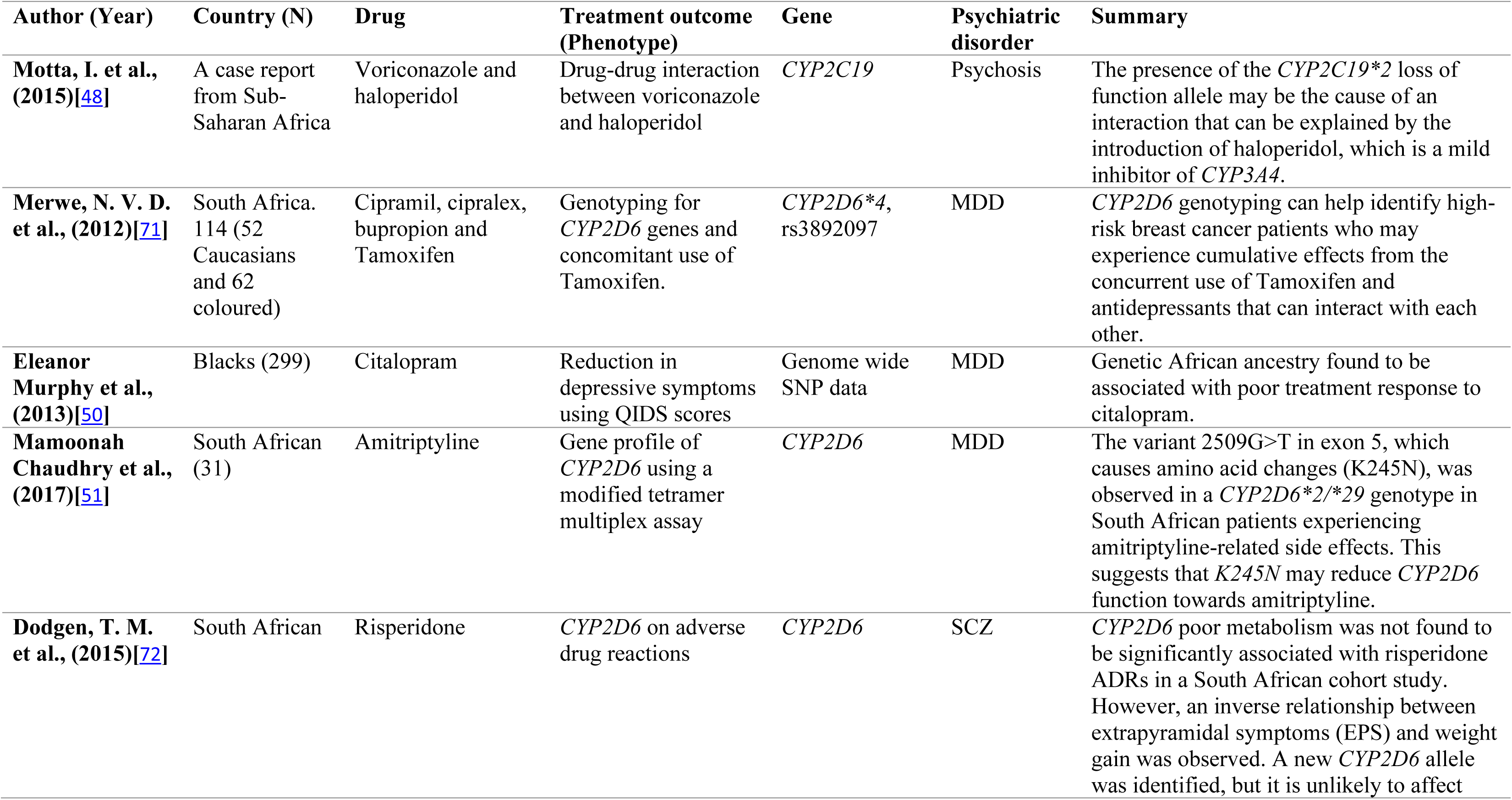
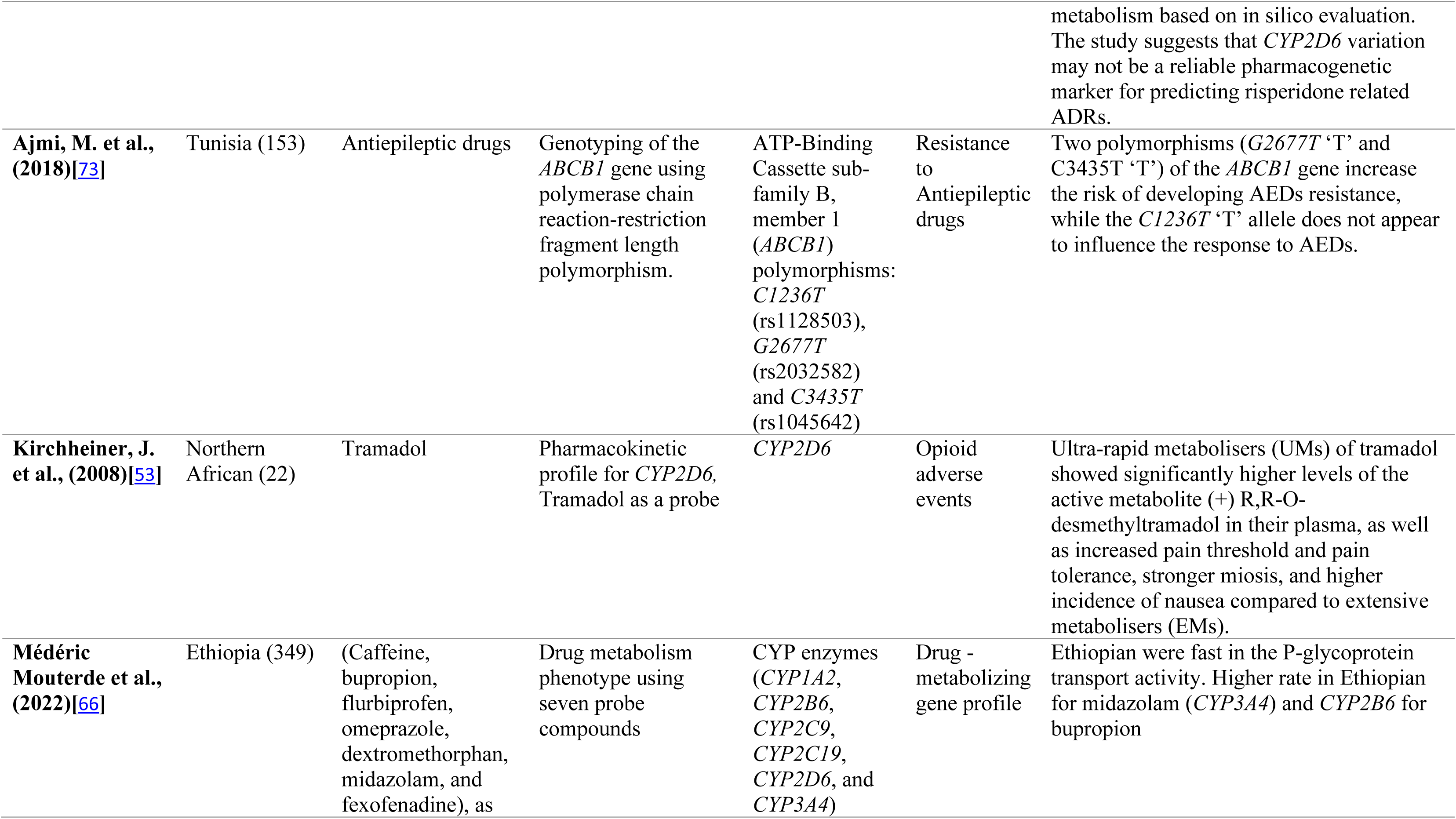
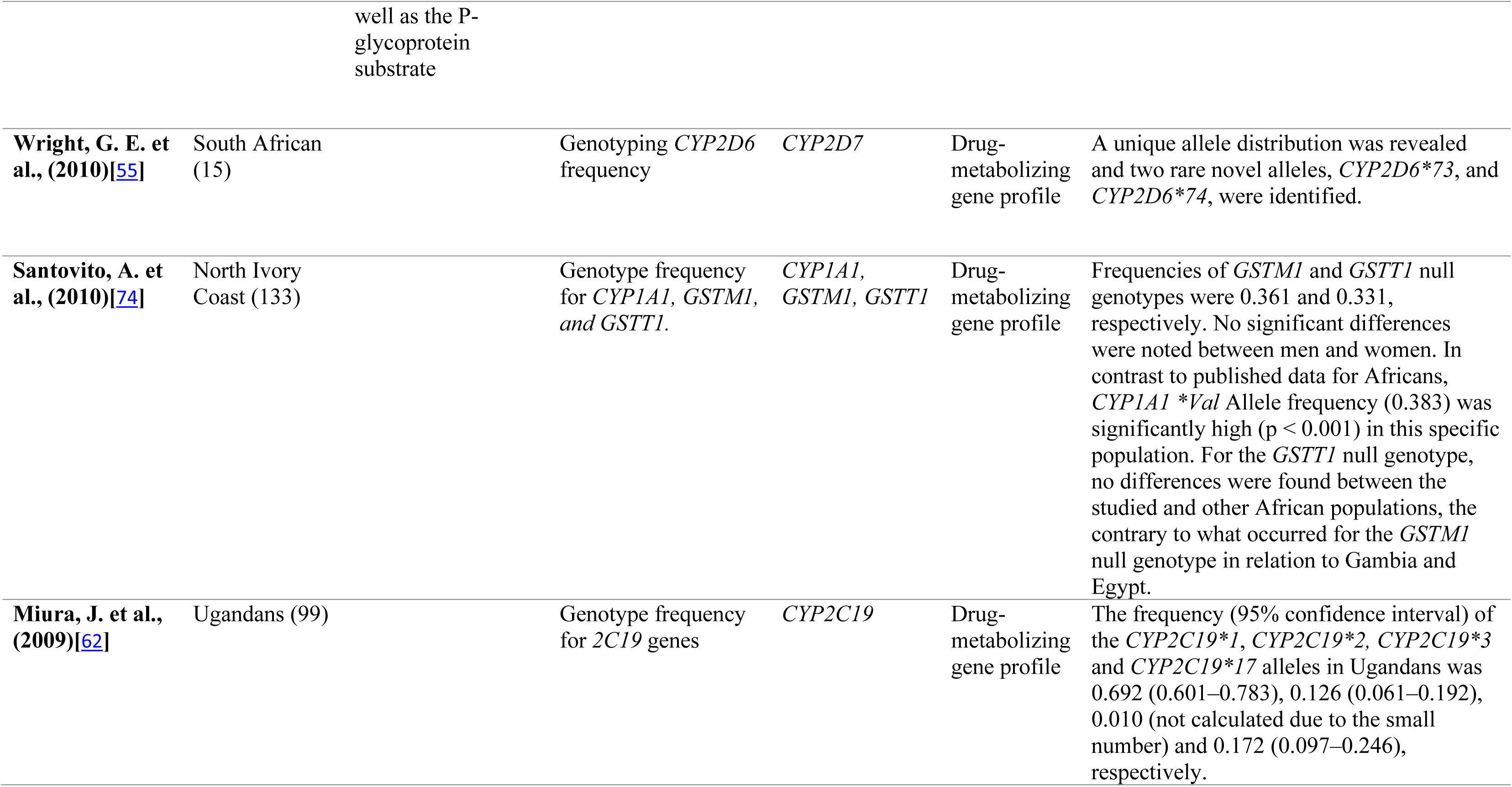
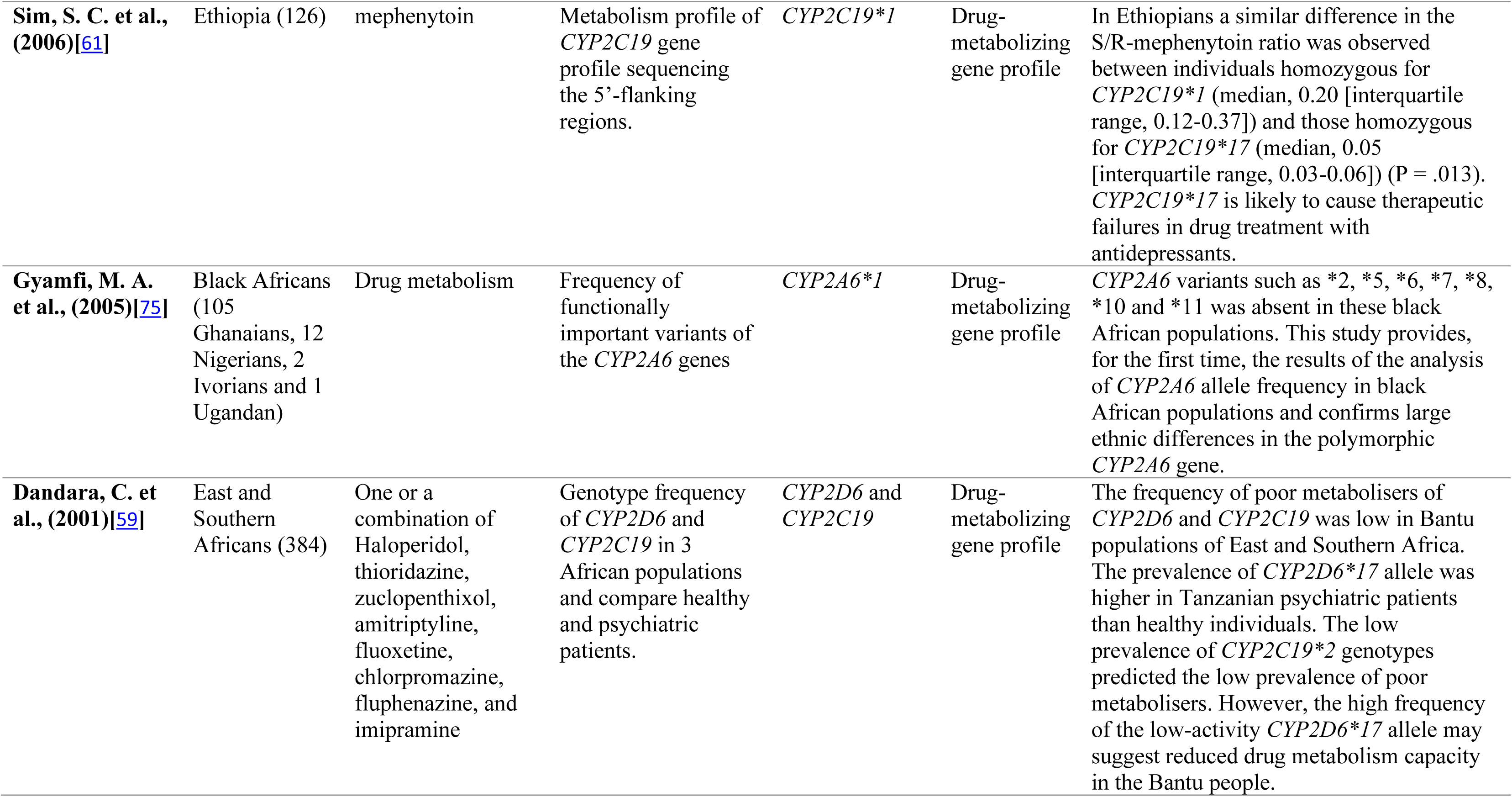
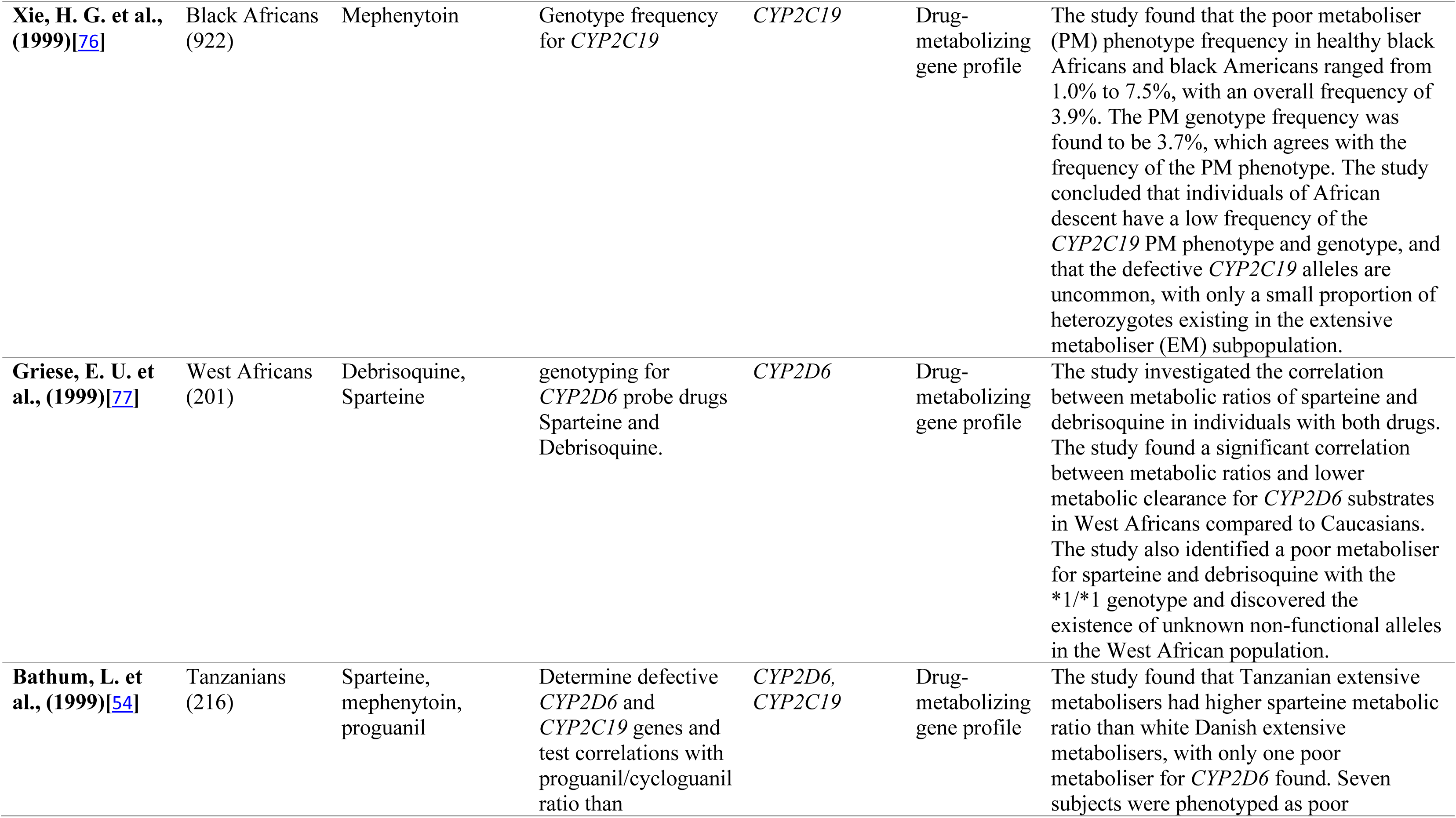
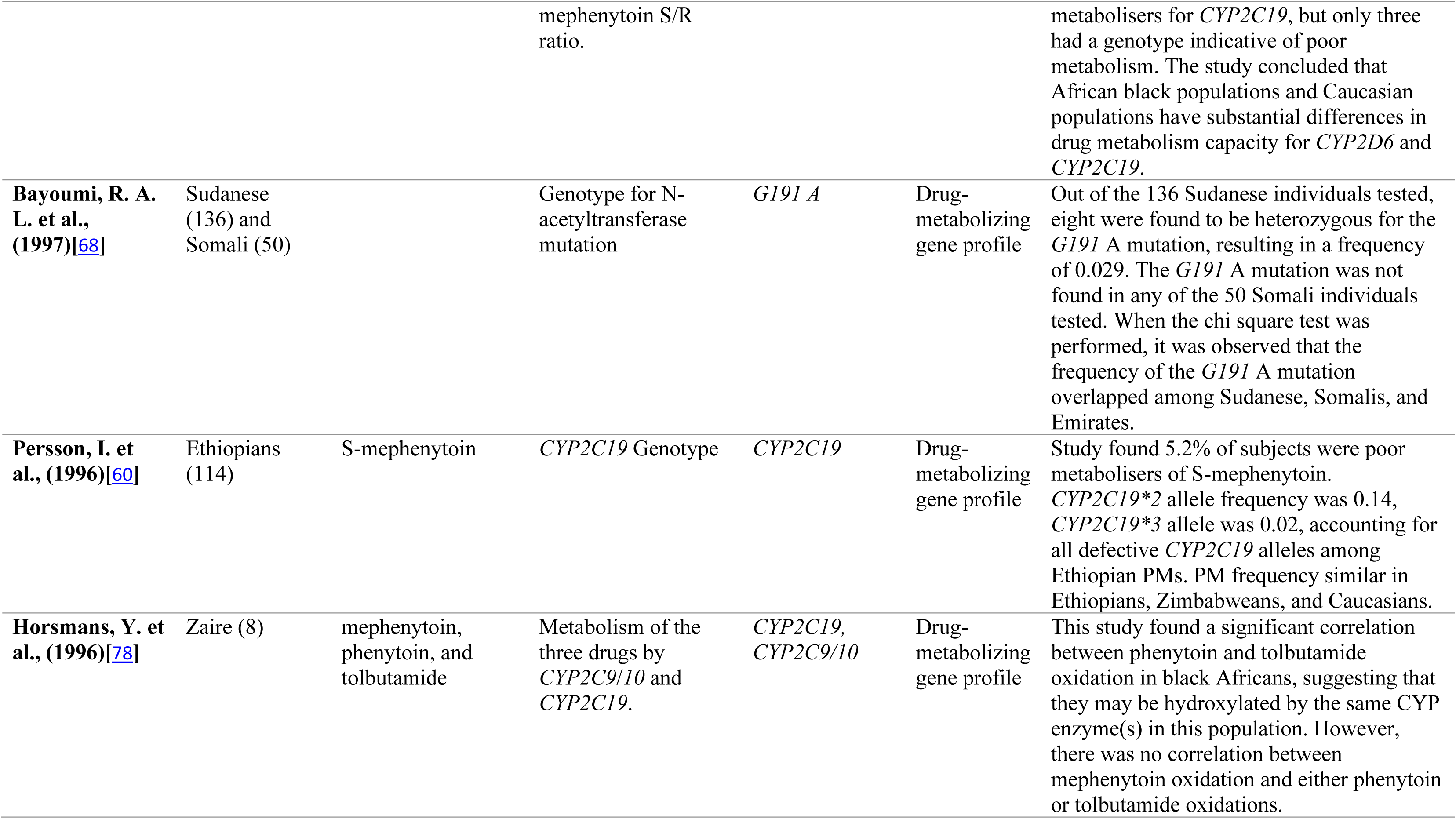
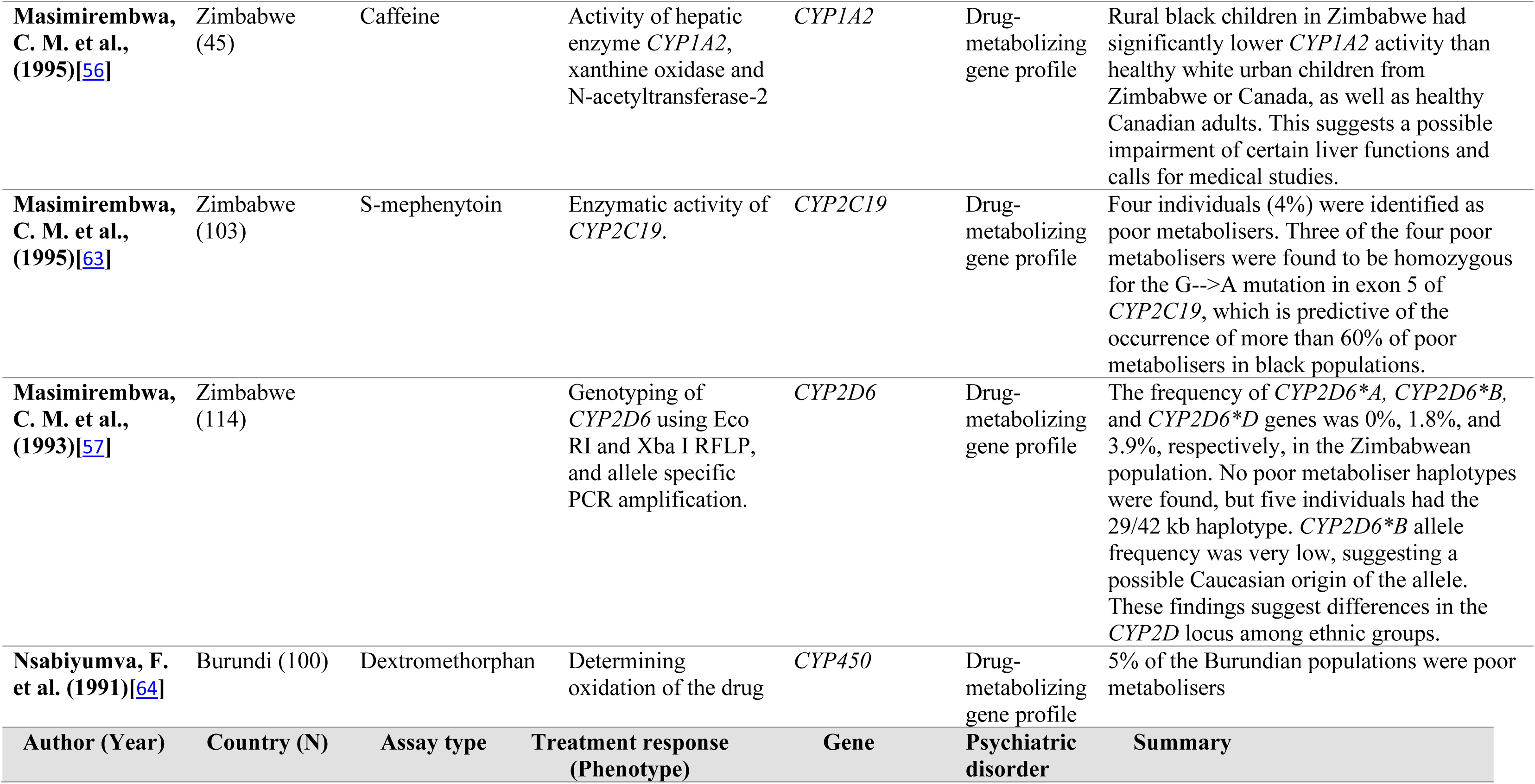
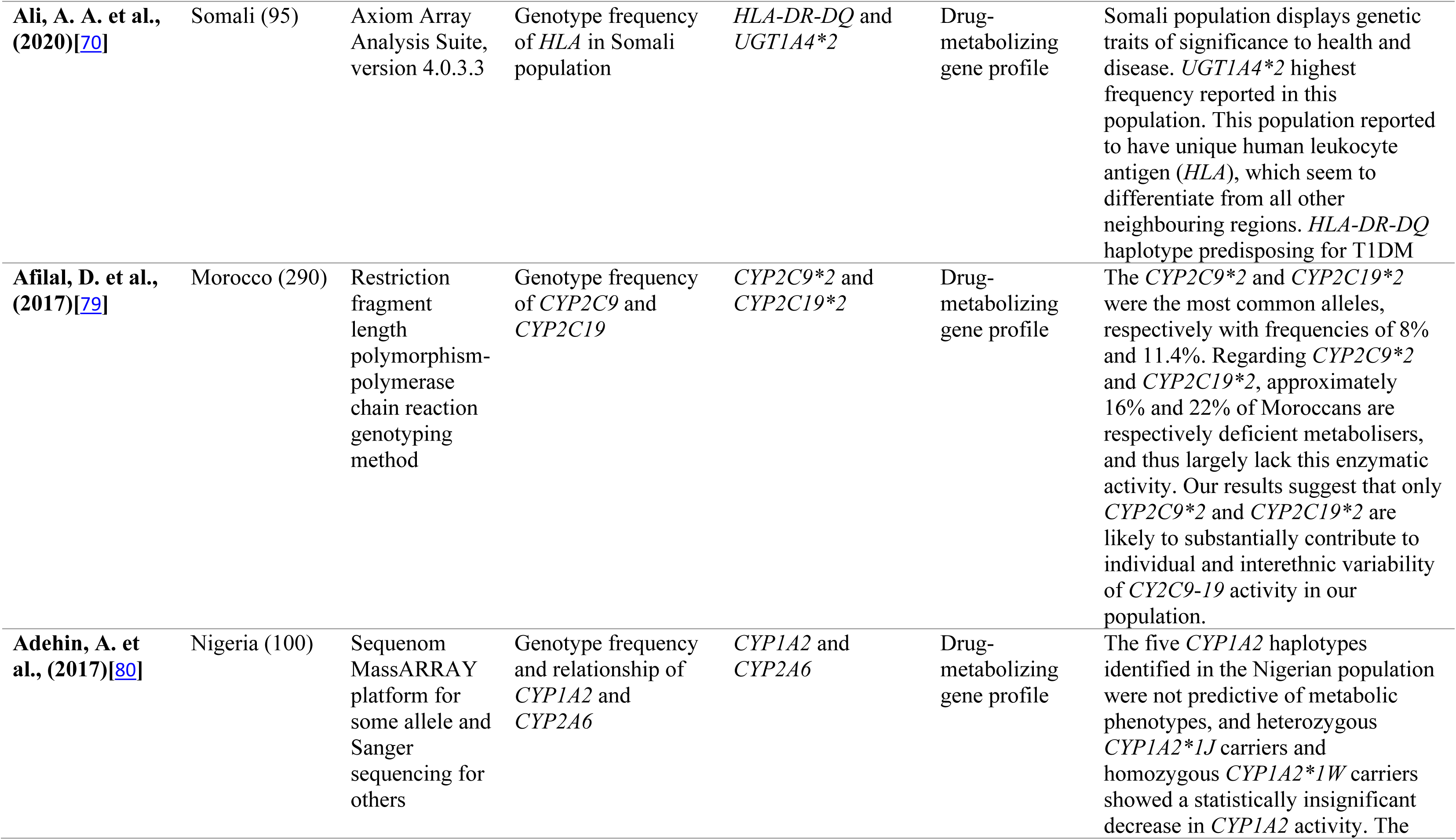
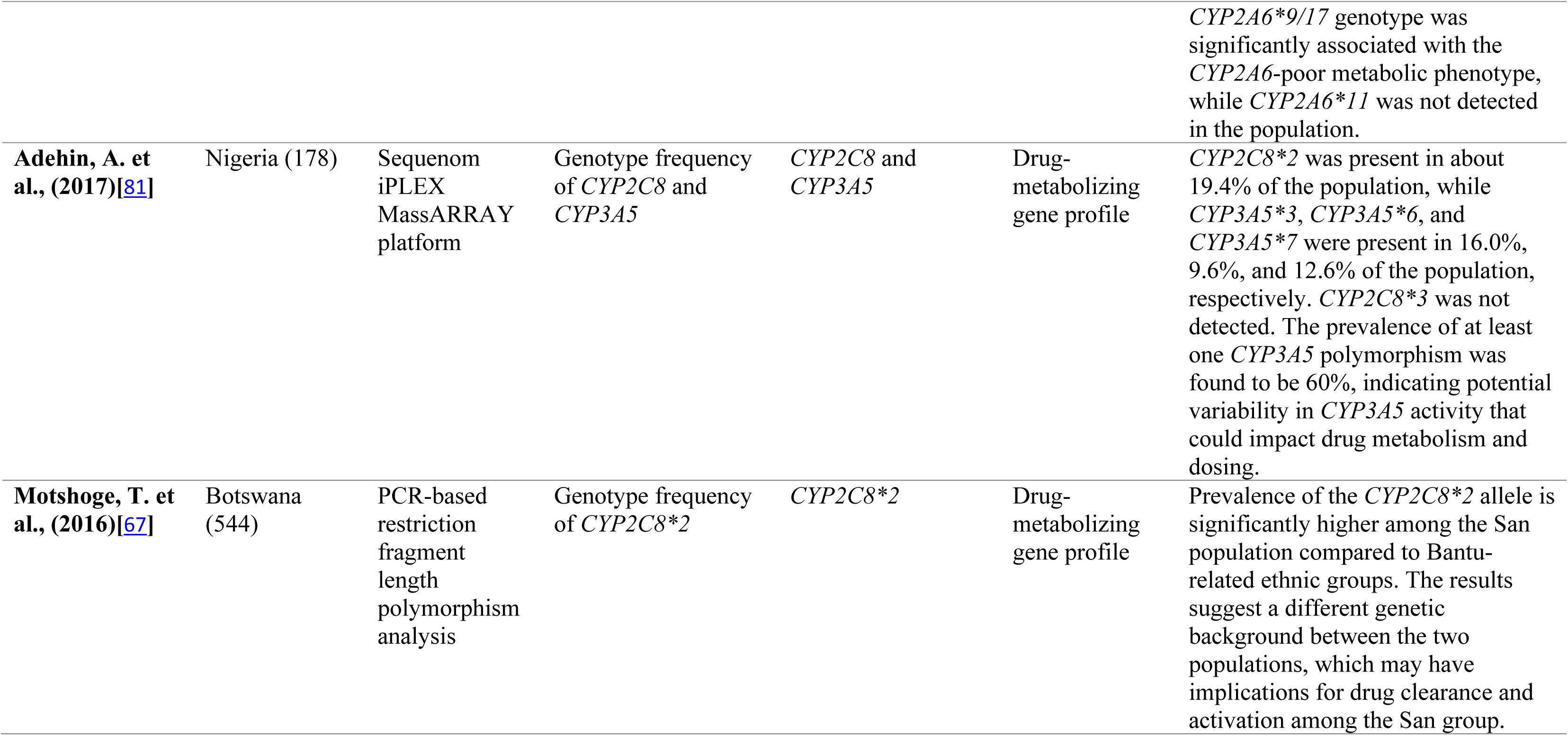
Pharmacokinetic and genetic profile among individuals of African ancestry.

| Author (Year) | Country (N) | Drug | Treatment outcome (Phenotype) | Gene | Psychiatric disorder | Summary |
| --- | --- | --- | --- | --- | --- | --- |
| **Motta, I. et al., (2015)[48]** | A case report from Sub-Saharan Africa | Voriconazole and haloperidol | Drug-drug interaction between voriconazole and haloperidol | *CYP2C19* | Psychosis | The presence of the *CYP2C19* *2 loss of function allele may be the cause of an interaction that can be explained by the introduction of haloperidol, which is a mild inhibitor of *CYP3A4*. |
| **Merwe, N. V. D. et al., (2012)[71]** | South Africa. 114 (52 Caucasians and 62 coloured) | Cipramil, cipralex, bupropion and Tamoxifen | Genotyping for *CYP2D6* genes and concomitant use of Tamoxifen. | *CYP2D6* *4, rs3892097 | MDD | *CYP2D6* genotyping can help identify high-risk breast cancer patients who may experience cumulative effects from the concurrent use of Tamoxifen and antidepressants that can interact with each other. |
| **Eleanor Murphy et al., (2013)[50]** | Blacks (299) | Citalopram | Reduction in depressive symptoms using QIDS scores | Genome wide SNP data | MDD | Genetic African ancestry found to be associated with poor treatment response to citalopram. |
| **Mamoonah Chaudhry et al., (2017)[51]** | South African (31) | Amitriptyline | Gene profile of *CYP2D6* using a modified tetramer multiplex assay | *CYP2D6* | MDD | The variant 2509G>T in exon 5, which causes amino acid changes (K245N), was observed in a *CYP2D6* *2/*29 genotype in South African patients experiencing amitriptyline-related side effects. This suggests that *K245N* may reduce *CYP2D6* function towards amitriptyline. |
| **Dodgen, T. M. et al., (2015)[72]** | South African | Risperidone | *CYP2D6* on adverse drug reactions | *CYP2D6* | SCZ | *CYP2D6* poor metabolism was not found to be significantly associated with risperidone ADRs in a South African cohort study. However, an inverse relationship between extrapyramidal symptoms (EPS) and weight gain was observed. A new *CYP2D6* allele was identified, but it is unlikely to affect metabolism based on in silico evaluation. The study suggests that *CYP2D6* variation may not be a reliable pharmacogenetic marker for predicting risperidone related ADRs. |
| **Ajmi, M. et al., (2018)**[73] | Tunisia (153) | Antiepileptic drugs | Genotyping of the *ABCB1* gene using polymerase chain reaction-restriction fragment length polymorphism. | ATP-Binding Cassette sub-family B, member 1 (*ABCB1*) polymorphisms: *C1236T* (rs1128503), *G2677T* (rs2032582) and *C3435T* (rs1045642) | Resistance to Antiepileptic drugs | Two polymorphisms (*G2677T* ‘T’ and *C3435T* ‘T’) of the *ABCB1* gene increase the risk of developing AEDs resistance, while the *C1236T* ‘T’ allele does not appear to influence the response to AEDs. |
| **Kirchheiner, J. et al., (2008)**[53] | Northern African (22) | Tramadol | Pharmacokinetic profile for *CYP2D6*, Tramadol as a probe | *CYP2D6* | Opioid adverse events | Ultra-rapid metabolisers (UMs) of tramadol showed significantly higher levels of the active metabolite (+) R,R-O-desmethyltramadol in their plasma, as well as increased pain threshold and pain tolerance, stronger miosis, and higher incidence of nausea compared to extensive metabolisers (EMs). |
| **Médéric Mouterde et al., (2022)**[66] | Ethiopia (349) | (Caffeine, bupropion, flurbiprofen, omeprazole, dextromethorphan, midazolam, and fexofenadine), as well as the P-glycoprotein substrate | Drug metabolism phenotype using seven probe compounds | CYP enzymes (*CYP1A2*, *CYP2B6*, *CYP2C9*, *CYP2C19*, *CYP2D6*, and *CYP3A4*) | Drug - metabolizing gene profile | Ethiopian were fast in the P-glycoprotein transport activity. Higher rate in Ethiopian for midazolam (*CYP3A4*) and *CYP2B6* for bupropion |
| **Wright, G. E. et al., (2010)**[55] | South African (15) |  | Genotyping *CYP2D6* frequency | *CYP2D7* | Drug-metabolizing gene profile | A unique allele distribution was revealed and two rare novel alleles, *CYP2D6\*73*, and *CYP2D6\*74*, were identified. |
| **Santovito, A. et al., (2010)**[74] | North Ivory Coast (133) | | Genotype frequency for *CYP1A1*, *GSTM1*, and *GSTT1*. | *CYP1A1*, *GSTM1*, *GSTT1* | Drug-metabolizing gene profile | Frequencies of *GSTM1* and *GSTT1* null genotypes were 0.361 and 0.331, respectively. No significant differences were noted between men and women. In contrast to published data for Africans, *CYP1A1* * *Val* Allele frequency (0.383) was significantly high ( $p < 0.001$ ) in this specific population. For the *GSTT1* null genotype, no differences were found between the studied and other African populations, the contrary to what occurred for the *GSTM1* null genotype in relation to Gambia and Egypt. |
| **Miura, J. et al., (2009)**[62] | Ugandans (99) |  | Genotype frequency for *2C19* genes | *CYP2C19* | Drug-metabolizing gene profile | The frequency (95% confidence interval) of the *CYP2C19\*1*, *CYP2C19\*2*, *CYP2C19\*3* and *CYP2C19\*17* alleles in Ugandans was 0.692 (0.601–0.783), 0.126 (0.061–0.192), 0.010 (not calculated due to the small number) and 0.172 (0.097–0.246), respectively. |
| **Sim, S. C. et al., (2006)**[61] | Ethiopia (126) | mephenytoin | Metabolism profile of *CYP2C19* gene profile sequencing the 5'-flanking regions. | *CYP2C19\*1* | Drug-metabolizing gene profile | In Ethiopians a similar difference in the S/R-mephenytoin ratio was observed between individuals homozygous for *CYP2C19\*1* (median, 0.20 [interquartile range, 0.12-0.37]) and those homozygous for *CYP2C19\*17* (median, 0.05 [interquartile range, 0.03-0.06]) (P = .013). *CYP2C19\*17* is likely to cause therapeutic failures in drug treatment with antidepressants. |
| **Gyamfi, M. A. et al., (2005)**[75] | Black Africans (105<br>Ghanaians, 12<br>Nigerians, 2<br>Ivorians and 1<br>Ugandan) | Drug metabolism | Frequency of functionally important variants of the *CYP2A6* genes | *CYP2A6\*1* | Drug-metabolizing gene profile | *CYP2A6* variants such as *2, *5, *6, *7, *8, *10 and *11 was absent in these black African populations. This study provides, for the first time, the results of the analysis of *CYP2A6* allele frequency in black African populations and confirms large ethnic differences in the polymorphic *CYP2A6* gene. |
| **Dandara, C. et al., (2001)**[59] | East and Southern Africans (384) | One or a combination of Haloperidol, thioridazine, zuclopenthixol, amitriptyline, fluoxetine, chlorpromazine, fluphenazine, and imipramine | Genotype frequency of *CYP2D6* and *CYP2C19* in 3 African populations and compare healthy and psychiatric patients. | *CYP2D6* and *CYP2C19* | Drug-metabolizing gene profile | The frequency of poor metabolisers of *CYP2D6* and *CYP2C19* was low in Bantu populations of East and Southern Africa. The prevalence of *CYP2D6\*17* allele was higher in Tanzanian psychiatric patients than healthy individuals. The low prevalence of *CYP2C19\*2* genotypes predicted the low prevalence of poor metabolisers. However, the high frequency of the low-activity *CYP2D6\*17* allele may suggest reduced drug metabolism capacity in the Bantu people. |
| **Xie, H. G. et al., (1999)[76]** | Black Africans (922) | Mephenytoin | Genotype frequency for *CYP2C19* | *CYP2C19* | Drug-metabolizing gene profile | The study found that the poor metaboliser (PM) phenotype frequency in healthy black Africans and black Americans ranged from 1.0% to 7.5%, with an overall frequency of 3.9%. The PM genotype frequency was found to be 3.7%, which agrees with the frequency of the PM phenotype. The study concluded that individuals of African descent have a low frequency of the *CYP2C19* PM phenotype and genotype, and that the defective *CYP2C19* alleles are uncommon, with only a small proportion of heterozygotes existing in the extensive metaboliser (EM) subpopulation. |
| **Griese, E. U. et al., (1999)[77]** | West Africans (201) | Debrisoquine, Sparteine | genotyping for *CYP2D6* probe drugs Sparteine and Debrisoquine. | *CYP2D6* | Drug-metabolizing gene profile | The study investigated the correlation between metabolic ratios of sparteine and debrisoquine in individuals with both drugs. The study found a significant correlation between metabolic ratios and lower metabolic clearance for *CYP2D6* substrates in West Africans compared to Caucasians. The study also identified a poor metaboliser for sparteine and debrisoquine with the *1/*1 genotype and discovered the existence of unknown non-functional alleles in the West African population. |
| **Bathum, L. et al., (1999)[54]** | Tanzanians (216) | Sparteine, mephenytoin, proguanil | Determine defective *CYP2D6* and *CYP2C19* genes and test correlations with proguanil/cycloguanil ratio than | *CYP2D6*, *CYP2C19* | Drug-metabolizing gene profile | The study found that Tanzanian extensive metabolisers had higher sparteine metabolic ratio than white Danish extensive metabolisers, with only one poor metaboliser for *CYP2D6* found. Seven subjects were phenotyped as poor |
|  |  |  | mephenytoin S/R ratio. |  |  | metabolisers for *CYP2C19*, but only three had a genotype indicative of poor metabolism. The study concluded that African black populations and Caucasian populations have substantial differences in drug metabolism capacity for *CYP2D6* and *CYP2C19*. |
| **Bayoumi, R. A. L. et al., (1997)[68]** | Sudanese (136) and Somali (50) |  | Genotype for N-acetyltransferase mutation | *G191 A* | Drug-metabolizing gene profile | Out of the 136 Sudanese individuals tested, eight were found to be heterozygous for the *G191 A* mutation, resulting in a frequency of 0.029. The *G191 A* mutation was not found in any of the 50 Somali individuals tested. When the chi square test was performed, it was observed that the frequency of the *G191 A* mutation overlapped among Sudanese, Somalis, and Emirates. |
| **Persson, I. et al., (1996)[60]** | Ethiopians (114) | S-mephenytoin | *CYP2C19* Genotype | *CYP2C19* | Drug-metabolizing gene profile | Study found 5.2% of subjects were poor metabolisers of S-mephenytoin. *CYP2C19* *2 allele frequency was 0.14, *CYP2C19* *3 allele was 0.02, accounting for all defective *CYP2C19* alleles among Ethiopian PMs. PM frequency similar in Ethiopians, Zimbabweans, and Caucasians. |
| **Horsmans, Y. et al., (1996)[78]** | Zaire (8) | mephenytoin, phenytoin, and tolbutamide | Metabolism of the three drugs by *CYP2C9/10* and *CYP2C19*. | *CYP2C19*, *CYP2C9/10* | Drug-metabolizing gene profile | This study found a significant correlation between phenytoin and tolbutamide oxidation in black Africans, suggesting that they may be hydroxylated by the same CYP enzyme(s) in this population. However, there was no correlation between mephenytoin oxidation and either phenytoin or tolbutamide oxidations. |

| **Masimirembwa, C. M. et al., (1995)[56]** | Zimbabwe (45) | Caffeine | Activity of hepatic enzyme *CYP1A2*, xanthine oxidase and N-acetyltransferase-2 | *CYP1A2* | Drug-metabolizing gene profile | Rural black children in Zimbabwe had significantly lower *CYP1A2* activity than healthy white urban children from Zimbabwe or Canada, as well as healthy Canadian adults. This suggests a possible impairment of certain liver functions and calls for medical studies. |
| --- | --- | --- | --- | --- | --- | --- |
| **Masimirembwa, C. M. et al., (1995)[63]** | Zimbabwe (103) | S-mephenytoin | Enzymatic activity of *CYP2C19*. | *CYP2C19* | Drug-metabolizing gene profile | Four individuals (4%) were identified as poor metabolisers. Three of the four poor metabolisers were found to be homozygous for the G-->A mutation in exon 5 of *CYP2C19*, which is predictive of the occurrence of more than 60% of poor metabolisers in black populations. |
| **Masimirembwa, C. M. et al., (1993)[57]** | Zimbabwe (114) |  | Genotyping of *CYP2D6* using Eco RI and Xba I RFLP, and allele specific PCR amplification. | *CYP2D6* | Drug-metabolizing gene profile | The frequency of *CYP2D6* *A, *CYP2D6* *B, and *CYP2D6* *D genes was 0%, 1.8%, and 3.9%, respectively, in the Zimbabwean population. No poor metaboliser haplotypes were found, but five individuals had the 29/42 kb haplotype. *CYP2D6* *B allele frequency was very low, suggesting a possible Caucasian origin of the allele. These findings suggest differences in the *CYP2D* locus among ethnic groups. |
| **Nsabiyumva, F. et al. (1991)[64]** | Burundi (100) | Dextromethorphan | Determining oxidation of the drug | *CYP450* | Drug-metabolizing gene profile | 5% of the Burundian populations were poor metabolisers |
| Author (Year) | Country (N) | Assay type | Treatment response (Phenotype) | Gene | Psychiatric disorder | Summary |
| **Ali, A. A. et al., (2020)**[70] | Somali (95) | Axiom Array Analysis Suite, version 4.0.3.3 | Genotype frequency of *HLA* in Somali population | *HLA-DR-DQ* and *UGT1A4\*2* | Drug-metabolizing gene profile | Somali population displays genetic traits of significance to health and disease. *UGT1A4\*2* highest frequency reported in this population. This population reported to have unique human leukocyte antigen (*HLA*), which seem to differentiate from all other neighbouring regions. *HLA-DR-DQ* haplotype predisposing for T1DM |
| **Afilal, D. et al., (2017)**[79] | Morocco (290) | Restriction fragment length polymorphism-polymerase chain reaction genotyping method | Genotype frequency of *CYP2C9* and *CYP2C19* | *CYP2C9\*2* and *CYP2C19\*2* | Drug-metabolizing gene profile | The *CYP2C9\*2* and *CYP2C19\*2* were the most common alleles, respectively with frequencies of 8% and 11.4%. Regarding *CYP2C9\*2* and *CYP2C19\*2*, approximately 16% and 22% of Moroccans are respectively deficient metabolisers, and thus largely lack this enzymatic activity. Our results suggest that only *CYP2C9\*2* and *CYP2C19\*2* are likely to substantially contribute to individual and interethnic variability of *CYP2C9-19* activity in our population. |
| **Adehin, A. et al., (2017)**[80] | Nigeria (100) | Sequenom MassARRAY platform for some allele and Sanger sequencing for others | Genotype frequency and relationship of *CYP1A2* and *CYP2A6* | *CYP1A2* and *CYP2A6* | Drug-metabolizing gene profile | The five *CYP1A2* haplotypes identified in the Nigerian population were not predictive of metabolic phenotypes, and heterozygous *CYP1A2\*1J* carriers and homozygous *CYP1A2\*1W* carriers showed a statistically insignificant decrease in *CYP1A2* activity. The |
|  |  |  |  |  |  | *CYP2A6\*9/17* genotype was significantly associated with the *CYP2A6*-poor metabolic phenotype, while *CYP2A6\*11* was not detected in the population. |
| **Adehin, A. et al., (2017)**[81] | Nigeria (178) | Sequenom iPLEX MassARRAY platform | Genotype frequency of *CYP2C8* and *CYP3A5* | *CYP2C8* and *CYP3A5* | Drug-metabolizing gene profile | *CYP2C8\*2* was present in about 19.4% of the population, while *CYP3A5\*3*, *CYP3A5\*6*, and *CYP3A5\*7* were present in 16.0%, 9.6%, and 12.6% of the population, respectively. *CYP2C8\*3* was not detected. The prevalence of at least one *CYP3A5* polymorphism was found to be 60%, indicating potential variability in *CYP3A5* activity that could impact drug metabolism and dosing. |
| **Motshoge, T. et al., (2016)**[67] | Botswana (544) | PCR-based restriction fragment length polymorphism analysis | Genotype frequency of *CYP2C8\*2* | *CYP2C8\*2* | Drug-metabolizing gene profile | Prevalence of the *CYP2C8\*2* allele is significantly higher among the San population compared to Bantu-related ethnic groups. The results suggest a different genetic background between the two populations, which may have implications for drug clearance and activation among the San group. |

The other pharmacogene of interest in African population was the *CYP2C19* gene that is known to influence the metabolism of a large number of clinically relevant drugs and drug classes such as antidepressants, benzodiazepines, mephenytoin, proton pump inhibitors (PPIs) [58]. A study using samples from South Africa (Venda), Tanzania, and Zimbabwe reported the low prevalence of poor metabolisers determined by the frequencies of *CYP2C19*2* genotypes[59]. No significant difference was observed in the frequency of *CYP2D6* or *CYP2C19* poor metabolisers genotypes between psychiatric patients and healthy individuals [59]. A study that examined the *CYP2C19* profile on 140 Ethiopians found that 5.2% of them were poor metabolisers of S-mephenytoin, with frequency of 14% for *CYP2C19*2* allele and 2% for *CYP2C19*3* allele [60]. Among the poor metabolisers, three were homozygous for *CYP2C19*2*, and the remaining three were heterozygous for both *CYP2C19*2* and *CYP2C19*3*. Furthermore, the *CYP2C19*2* and *CYP2C19*3* alleles accounted for all defective *CYP2C19* alleles among the Ethiopian poor metabolisers [60]. Ethiopians individuals homozygous for the *CYP2C19*1* allele had a different S/R-mephenytoin ratio than those homozygous for *CYP2C19*17* with potential implication of this variant to antidepressants treatment [61]. In Ugandans, the frequency of the *CYP2C19*\*1, *CYP2C19*\*2, *CYP2C19*\*3, and *CYP2C19*\*17 alleles was 69.2%, 12.6%, 1.0%, and 17.2%, respectively [62]. A study that investigated the activity of *CYP2C19* in 103 Zimbabwean adults also found that four of the subjects were poor metabolizes[63]. Among 84 subjects phenotyped, a mutation in exon 5 of *CYP2C19* causes a cryptic splicing site that results in non-functional proteins. Three of the four poor metabolisers were homozygous for the mutation in exon 5 of *CYP2C19*, but one was heterozygous, suggesting that exon 5 mutations may be predictive of more than 60% of black poor metabolisers [63]. In a study in Burundi, it was reported that 5% of the population were classified as poor metabolisers [64].

*CYP3A4* was another important pharmacogene, together with *CYP3A5* account for 30% of the hepatic cytochrome P450, and half of medications that are oxidatively metabolised by liver are *CYP3A* substrates [65]. Ethiopians have a fast P-glycoprotein transport activity, with a higher rate for midazolam (*CYP3A4*) and bupropion *(CYP2B6)* [66]. On the other hand, a study comparing two ethnic groups in Botswana showed a significantly higher prevalence of the *CYP2C8*\*2 allele in the San (or Bushmen) than the Bantu-related ethnic groups [67].

A study comparing the activity of *CYP1A2* in black Zimbabwean children with Canadian adults found a significantly lower indexes of *CYP1A2* activity than that of healthy white urban children from Zimbabwe or healthy Canadian adults [56]. In a study of 136 Sudanese individuals, eight of them were heterozygous for the *G191A* mutation[68]. However, none of the 50 Somali individuals tested carried the *G191A* mutation [68]. The Human Leukocyte Antigen (HLA) alleles were also examined in African population for their role in the pharmacogenetics of drug hypersensitivity [69] and studies found both high frequency and unique HLA characteristics such as *HLA-DR-DQ* haplotypes [70].

## Discussion

This article presents a systematic review of the status of psychiatric pharmacogenomics in African populations, highlighting the challenges and opportunities for setting up PGx testing and expanding future research efforts in the region. The high genetic variability of African populations is well-documented [82], and this variability has implications on the way drugs are metabolized and affects treatment effectiveness in these populations. By studying the unique genetic characteristics of African populations, researchers have attempted to present novel insights into how the effectiveness of drugs can be influenced by genetic variations [29, 36]. These findings offer a promising opportunity to advance pharmacogenomics research and improve health outcomes for not only Africans but also individuals of all races and ethnicities around the world [36].

In Africa, pharmacogenomic research is evolving very slowly and the number of published studies on psychiatric pharmacogenomics is relatively low; as such, the translation of these findings into clinical practice is very limited compared to other regions of the world [83]. Within African regions, there is a significant discrepancy in the number of studies on psychiatric pharmacogenomics, with a higher publications observed in eastern and southern African countries, particularly in South Africa and Zimbabwe, when compared to other African regions [39–42, 51, 57]. The regional variations in research output may be attributed to several factors, including discrepancies in access to research funding, the presence of adequate infrastructure and expertise, as well as sociocultural and ethical considerations influencing the feasibility and prioritization of genomic research activities.

In this review, we assessed 42 pharmacogenomic studies that explored the impact of genetic factors on drug response, remission rates, and drug metabolism in individuals diagnosed with major psychiatric disorders, including SCZ, MDD, and BD. Studies involving individuals with SCZ have identified genetic variations within genes such as *CYP2C9*, *CYP2C19*, *MYO7B*, *MTRR*, *RGS4*, *CYP1A2*, and *CYP1A*21F* associated with treatment outcomes and influencing drug metabolism [41, 42, 84]. For instance, the CLOZUK study found that individuals of sub-Saharan African ancestry metabolize clozapine faster than people of European descent [37], which has significant implications for adjusting dosing and improve treatment efficacy of this drug. CYP enzymes, such as *CYP2D6*, *CYP1A2*, and *CYP3A4/5,* play a crucial role in the metabolism of antipsychotic medications, and the polymorphic alleles of these proteins are associated with altered plasma levels of these medications [85], impacting the potential for developing adverse reactions and therapeutic efficacy. It is also well established that an estimated 40% of antipsychotics are metabolized by *CYP2D6*, 23% by *CYP3A4*, and 18% by *CYP1A* [86], making PGx testing useful in guiding drug therapy for better treatment outcomes for psychiatric patients.

The scope of this review extends to pharmacogenomic studies exploring various candidate genes that have been implicated in influencing susceptibility to adverse drug reactions (ADR), a harmful or unpleasant reaction resulting from an intervention related to the use of a medicinal product [87]. The inclusion of ADR as part of drug response criteria is of paramount importance in assessing the efficacy and safety of medications. One such example from this review is the *CYP2D6*2/*29* genotype, where the variant 2509G>T causes amitriptyline treatment-related side effects in South African patients, indicating the relevance of genotyping such variants to identify patients at high risk of ADR [51].

Significant differences in the frequencies of genetic variations involved in drug metabolism among psychiatric patients in Tanzania and South Africa has provided valuable insights into the pharmacogenetics of cytochrome enzymes in African populations. For example, the Bantu people of East and Southern Africa have a low prevalence of the deficient *CYP2D6* and *CYP2C19* activity, but a high frequency of the low-activity *CYP2D6*17* allele [59]. A study conducted in Botswana comparing the prevalence of the *CYP2C8*2* allele found a significantly higher prevalence of this allele in the San group than the Bantu-related ethnic groups [67]. Another study found that rural black children in Zimbabwe had lower indexes of *CYP1A2* activity than healthy white urban children from Zimbabwe or healthy Canadians [56]. The study conducted on Zimbabwean adults regarding the *CYP2C19* gene revealed that the mutation in exon 5 of *CYP2C19* resulted in a non-functional protein among adults of poor metabolisers [63]. As many drugs used in psychiatry are metabolized by *CYP2D6* and *CYP2C19* enzymes [88], this finding has clinical implications to address ADRs through understanding their impact on drug metabolism, helping in decision-making to optimize the treatment of patients of African origin Despite the limited evidence in African populations [90], PGx testing has shown significant benefit in white Europeans [91] and Asians [92]. Studies and clinical reports have demonstrated the benefits of genetic-guided therapy for psychiatric patients. For instance, a five-gene [*CYP2D6*, *CYP2C19*, *CYP1A2*, *SLC6A4*, and *HTR2A*] PGx testing by Mayo Clinic (in the United States) resulted in a significant increase in the number of responders after 8 weeks of antidepressant therapy compared to standard treatment [93]. In a Norwegian study involving 2,066 patients, *CYP2C19* ultra metabolisers and *CYP2C19* poor metabolisers were found to be more likely to switch escitalopram to another drug [94]. These examples indicate the importance of genetic-guided therapy across ancestries, including African populations. However, it may not be possible to directly translate existing PGx data into genetic testing for African populations since current PGx data originate in populations of European and Asian origin, while African populations are not well characterized. Moreover, differences in linkage disequilibrium (LD) and allele frequency pose challenges to the transferability of such data. For example, the genetic diversity of ADME genes is not uniform across African populations, particularly in terms of high-impact coding variation. The distinct differences between the Southern African population and the far Western African population as well as Europeans indicate that PGx strategies based on European variants may have limited applicability to African populations [95]. In a different disease context, functional variants in the *CYP2B6* gene contribute to a high risk of ADR to efavirenz in HIV patients in sub-Saharan Africa [96], and *CYP2D6* variations negatively influence the efficacy of primaquine [97, 98]. Studies included in this review provide a partial view of ADME variation in the African population and emphasize the importance of further study to understand the full range of variation. Precision medicine guidelines and PGx tools for African populations should take into account the unique ADME landscape found in African population.

Although few, existing studies in African populations underscore the importance of genetic variation in the treatment of patients with mental illness. For example, the *DRD2* gene variant predicts the response of African Americans to clozapine, indicating the need for genotype-guided drug prescribing in patients with similar genetic properties [99]. An additional example is the *GRM3* SNP-rs724226, which is associated with PANSS total score in African Americans who took risperidone [100]. One of the major reasons for the substantial differences in drug metabolism capacities between African black and Caucasian populations is the frequency of *CYP450* alleles, such as *CYP2D6*1* and *CYP2D6*4*, suggesting the need for a highly comprehensive pharmacogenomic testing platform [101, 102].

Studies on the application of pharmacogenomics-guided therapy in African psychiatric practice are insufficient, and there is a lack of standardized methods for conducting pharmacogenomic tests in African countries. This situation raises questions about the feasibility of applying pharmacogenomics-guided therapy in those countries. [36]. As has been shown in other populations, Africans could benefit from the use of PGx testing for monitoring psychiatric medication efficacy and safety. However, there are still gaps in the evidence base for psychiatric pharmacogenomics in African populations, as many candidate genes have proven difficult to replicate [103, 104]. Therefore, addressing these gaps through further research is essential, particularly in sub-Saharan countries where the burden of mental illness is high.

### Challenges and opportunities for pharmacogenomics in Africa

This review has revealed that the field of pharmacogenomics research and PGx in Africa is still in its early stages and is likely to encounter several challenges during its implementation [105]. *Firstly,* various factors such as insufficient financial support for research, inadequate infrastructure, and a shortage of trained staff, laboratories, and equipment to perform the tests could challenge to conduct research and implement PGx testing [106]. The limited funding available for pharmacogenomics research in Africa, coupled with the reluctance of ‘funding organizations’ to invest due to concerns regarding infrastructure and trained personnel, as well as the perceived complexity of genetic research in Africa, could lead to a vicious cycle where insufficient funding hampers both research capacity and clinical implementation of PGx testing. For example, funding constraints limit our ability to increase the diversity of global pharmacogenomic databases, which currently rely on data from European or Asian populations. As a result, the translation of existing pharmacogenomics knowledge to African populations could be limited, and the world may miss out on the potential benefits of utilizing the rare genomic diversity that exists only in African samples. Interestingly, there are some encouraging efforts underway to increase funding for pharmacogenomics research in Africa. For example, international organizations such as the NIH, Wellcome Trust, and the EU have a strong interest in providing short-term funding and other resources to support research in low- and middle-income countries, including in Africa [107]. *Secondly,* the lack of awareness among healthcare professionals, policymakers, and the general public presents a significant challenge to pharmacogenomics research in Africa. The absence of understanding the potential benefits of pharmacogenomics can restrict research funding, complicate clinical translation, and pose challenges to public engagement, thereby impeding the collection of essential genetic data. To address these challenges, efforts should be made to increase awareness through training programs, educational campaigns, and community outreach initiatives. It is also crucial to involve African researchers and clinicians in the development and implementation of pharmacogenomics initiatives to ensure that they are culturally appropriate and effective [108–111]. *Thirdly,* research ethics can be complex, and the research process may take longer than expected as it requires consideration of the diverse cultural and religious beliefs of the people ensuring that the research is conducted with respect and ethical principles. One way to achieve these goals is by involving local communities, which can help to ensure that the research is culturally appropriate, addresses community needs and concerns, and respects local values and beliefs [112, 113].

Moreover, the integration of electronic health records (EHR) in longitudinal cohorts may help to greatly increase sample size, to study uncommon exposures, rare treatment outcomes, and carry out comprehensive standardized analyses on drug response [114]. By incorporating diverse African populations into such cohorts, researchers can gain valuable insights into the genetic variations that impact drug metabolism and response [115]. This information guides the development of personalized treatment strategies and enhances medication safety and efficacy. Longitudinal cohorts also facilitate the study of environmental factors, socio-economic determinants, and lifestyle choices on drug response contributing to precision medicine initiatives to Africa’s genomic diversity [116]. Additionally, they enable the assessment of long-term medication effects and identification of rare or delayed adverse drug reactions specific to African populations [117]. Leveraging longitudinal cohorts strengthen Africa’s capacity for PGx research, inform clinical decision-making, and ultimately improve patient outcomes in precision medicine.

Despite the challenges, opportunities exist to advance pharmacogenomic research in Africa, especially with growing global interest in genomic diversity. National governments and international partners have shown strong commitment to supporting genomic research in developing countries, including in Africa. Initiatives such as the African Pharmacogenetics Consortium (APC) [118], Human Heredity and Health in Africa (H3Africa) [119] and Collaborative African Genomics Network (CAfGEN) [120] can accelerate pharmacogenomic research and implementation. Some African governments started to recognize the importance of pharmacogenomics for development and they encourage investment in research infrastructure and training programs [122, 123]. Pharmaceutical and biotechnology firms are also investing in Africa, focusing on drugs repurposing and development of new drugs [124, 125]. moreover, international collaborations between African and non-African research institutions contribute funding, expertise, and resources accelerating research in the continent [124].

In conclusion, this review highlights the significant pharmacogenomic diversity observed in individuals of African ancestry. While the number of studies is limited, existing evidence shows variability in the frequency, distribution, and type of genetic variants within drug-metabolizing enzymes and other candidate genes involved in the effectiveness of drugs in Africa. For example, *CYP2D6, CYP2C19, CYP3A4, CYP2B6, CYP2,* and *CYP1A2* vary widely across African populations. These genetic variations may result in altered drug concentrations, which can influence treatment outcomes, including efficacy and adverse reactions. Understanding the impact of these genetic variants on drug response is crucial for optimizing medication selection, dosing, and treatment outcomes in African populations. Much of the current pharmacogenomics knowledge in psychiatry is based on small-scale studies, emphasizing the need for increased funding and research infrastructure to enable the implementation of PGx testing in psychiatric treatment. To increase the representation of African populations in pharmacogenomic research and enhance our understanding of the genetic factors underlying patients’ response to pharmacological therapy, global consortia initiatives should expand access to next-generation sequencing facilities in the region. Further research into psychiatric pharmacogenomics and pharmacogenomic diversity has the potential to revolutionize scientific knowledge, but it requires a collaborative effort to close the knowledge gap and overcome implementation barriers.

## Data Availability

This is a systematic review and there is no individual data to share.

## References

1. Collaborators, G.B.D.M.D., Global, regional, and national burden of 12 mental disorders in 204 countries and territories, 1990-2019: a systematic analysis for the Global Burden of Disease Study 2019. Lancet Psychiatry, 2022. 9(2): p. 137–150.

2. Jorns-Presentati, A., et al., The prevalence of mental health problems in sub-Saharan adolescents: A systematic review. PLoS One, 2021. 16(5): p. e0251689.

3. Greene, M.C., et al., The epidemiology of psychiatric disorders in Africa: a scoping review. Lancet Psychiatry, 2021. 8(8): p. 717–731.

4. Kumar, S., et al., Antipsychotic Medication in Sub-Saharan Africa: A Systematic Literature Review. J Clin Psychopharmacol, 2020. 40(6): p. 541–552.

5. Entsuah, A.R., H. Huang, and M.E. Thase, Response and remission rates in different subpopulations with major depressive disorder administered venlafaxine, selective serotonin reuptake inhibitors, or placebo. Journal of Clinical Psychiatry, 2001. 62(11): p. 869–877.

6. Kozyra, M., M. Ingelman-Sundberg, and V.M. Lauschke, Rare genetic variants in cellular transporters, metabolic enzymes, and nuclear receptors can be important determinants of interindividual differences in drug response. Genet Med, 2017. 19(1): p. 20–29.

7. Investigators, G., M. Investigators, and S.D. Investigators, Common genetic variation and antidepressant efficacy in major depressive disorder: a meta-analysis of three genome-wide pharmacogenetic studies. Am J Psychiatry, 2013. 170(2): p. 207–17.

8. Pardinas, A.F., et al., Pharmacogenomic Variants and Drug Interactions Identified Through the Genetic Analysis of Clozapine Metabolism. Am J Psychiatry, 2019. 176(6): p. 477–486.

9. Quilty, L.C., et al., Dimensional personality traits and treatment outcome in patients with major depressive disorder. J Affect Disord, 2008. 108(3): p. 241–50.

10. Bulmash, E., et al., Personality, stressful life events, and treatment response in major depression. J Consult Clin Psychol, 2009. 77(6): p. 1067–77.

11. Bousman, C.A., P. Jaksa, and C. Pantelis, Systematic evaluation of commercial pharmacogenetic testing in psychiatry: a focus on CYP2D6 and CYP2C19 allele coverage and results reporting. Pharmacogenet Genomics, 2017. 27(11): p. 387–393.

12. Pardinas, A.F., M.J. Owen, and J.T.R. Walters, Pharmacogenomics: A road ahead for precision medicine in psychiatry. Neuron, 2021. 109(24): p. 3914–3929.

13. Kang, H.J., et al., Genetic Markers for Later Remission in Response to Early Improvement of Antidepressants. Int J Mol Sci, 2020. 21(14).

14. Lally, J., et al., Treatment-resistant schizophrenia: current insights on the pharmacogenomics of antipsychotics. Pharmgenomics Pers Med, 2016. 9: p. 117–129.

15. Jeiziner, C., et al., HLA-associated adverse drug reactions-scoping review. Clinical and Translational Science, 2021. 14(5): p. 1648–1658.

16. Crouch, D.J. and W.F. Bodmer, Polygenic inheritance, GWAS, polygenic risk scores, and the search for functional variants. Proceedings of the National Academy of Sciences, 2020. 117(32): p. 18924–18933.

17. Fagerness, J., et al., Pharmacogenetic-guided psychiatric intervention associated with increased adherence and cost savings. Am J Manag Care, 2014. 20(5): p. e146–56.

18. Lee, J.W., et al., The emerging era of pharmacogenomics: current successes, future potential, and challenges. Clinical genetics, 2014. 86(1): p. 21–28.

19. Lee, J.W., et al., The emerging era of pharmacogenomics: current successes, future potential, and challenges. Clin Genet, 2014. 86(1): p. 21–8.

20. Ripke, S., et al., Biological insights from 108 schizophrenia-associated genetic loci. Nature, 2014. 511(7510): p. 421-+.

21. Hou, L., et al., Genetic variants associated with response to lithium treatment in bipolar disorder: a genome-wide association study. The Lancet, 2016. 387(10023): p. 1085–1093.

22. International Consortium on Lithium, G., et al., Association of Polygenic Score for Schizophrenia and HLA Antigen and Inflammation Genes With Response to Lithium in Bipolar Affective Disorder: A Genome-Wide Association Study. JAMA Psychiatry, 2018. 75(1): p. 65–74.

23. Amare, A.T., et al., Association of polygenic score for major depression with response to lithium in patients with bipolar disorder. Mol Psychiatry, 2021. 26(6): p. 2457–2470.

24. Amare, A.T., et al., Association of polygenic score and the involvement of cholinergic and glutamatergic pathways with lithium treatment response in patients with bipolar disorder. Molecular Psychiatry, 2023.

25. Cearns, M., et al., Using polygenic scores and clinical data for bipolar disorder patient stratification and lithium response prediction: machine learning approach. Br J Psychiatry, 2022: p. 1–10.

26. Fabbri, C., Genetics in psychiatry: Methods, clinical applications and future perspectives. Psychiatry and Clinical Neurosciences Reports, 2022. 1(2).

27. Rayner, C., et al., Genetic influences on treatment-seeking for common mental health problems in the UK biobank. Behav Res Ther, 2019. 121: p. 103413.

28. Kohlrausch, F.B., et al., Naturalistic pharmacogenetic study of treatment resistance to typical neuroleptics in European-Brazilian schizophrenics. Pharmacogenet Genomics, 2008. 18(7): p. 599–609.

29. Rajman, I., et al., African Genetic Diversity: Implications for Cytochrome P450-mediated Drug Metabolism and Drug Development. EBioMedicine, 2017. 17: p. 67–74.

30. Martin, A.R., et al., The critical needs and challenges for genetic architecture studies in Africa. Curr Opin Genet Dev, 2018. 53: p. 113–120.

31. Omotoso, O.E., et al., Bridging the genomic data gap in Africa: implications for global disease burdens. Global Health, 2022. 18(1): p. 103.

32. Matimba, A., M. Dhoro, and C. Dandara, Is there a role of pharmacogenomics in Africa. Glob Health Epidemiol Genom, 2016. 1: p. e9.

33. Page, M.J., et al., The PRISMA 2020 statement: an updated guideline for reporting systematic reviews. BMJ, 2021. 372: p. n71.

34. Team, T.E., EndNote. 2013, Clarivate: Philadelphia, PA.

35. Amal Al, O. and D.J. Murry, Pharmacogenetics of the Cytochrome P450 Enzyme System: Review of Current Knowledge and Clinical Significance. Journal of Pharmacy Practice, 2016. 20(3): p. 206–218.

36. Radouani, F., et al., A review of clinical pharmacogenetics Studies in African populations. Per Med, 2020. 17(2): p. 155–170.

37. Pardinas, A.F., et al., Pharmacokinetics and pharmacogenomics of clozapine in an ancestrally diverse sample: a longitudinal analysis and genome-wide association study using UK clinical monitoring data. Lancet Psychiatry, 2023. 10(3): p. 209–219.

38. Campbell, D.B., et al., Ethnic stratification of the association of RGS4 variants with antipsychotic treatment response in schizophrenia. Biol Psychiatry, 2008. 63(1): p. 32–41.

39. Drogemoller, B.I., G.E.B. Wright, and L. Warnich, Considerations for rare variants in drug metabolism genes and the clinical implications. Expert Opinion on Drug Metabolism & Toxicology, 2014. 10(6): p. 873–884.

40. Drogemoller, B.I., et al., The identification of novel genetic variants associated with antipsychotic treatment response outcomes in first-episode schizophrenia patients. Pharmacogenetics and Genomics, 2016. 26(5): p. 235–242.

41. O’Connell, K.S., et al., The Potential Role of Regulatory Genes (DNMT3A, HDAC5, and HDAC9) in Antipsychotic Treatment Response in South African Schizophrenia Patients. Frontiers in Genetics, 2019. 10: p. 9.

42. Ovenden, E.S., et al., Fine-mapping of antipsychotic response genome-wide association studies reveals novel regulatory mechanisms. Pharmacogenomics, 2017. 18(2): p. 105–120.

43. Drogemoller, B.I., et al., Patterns of variation influencing antipsychotic treatment outcomes in South African first-episode schizophrenia patients. Pharmacogenomics, 2014. 15(2): p. 189–99.

44. Fijal, B.A., et al., Candidate-gene association analysis of response to risperidone in African-American and white patients with schizophrenia. Pharmacogenomics J, 2009. 9(5): p. 311–8.

45. Hwang, R., et al., Association study of 12 polymorphisms spanning the dopamine D(2) receptor gene and clozapine treatment response in two treatment refractory/intolerant populations. Psychopharmacology (Berl), 2005. 181(1): p. 179–87.

46. Ammar, H., et al., Clinical and genetic influencing factors on clozapine pharmacokinetics in Tunisian schizophrenic patients. Pharmacogenomics J, 2021. 21(5): p. 551–558.

47. Benmessaoud, D., et al., Excess of transmission of the G allele of the -1438A/G polymorphism of the 5-HT2A receptor gene in patients with schizophrenia responsive to antipsychotics. BMC Psychiatry, 2008. 8: p. 40.

48. Motta, I., et al., A probable drug-to-drug interaction between voriconazole and haloperidol in a slow metabolizer of CYP2C19. Infezioni in Medicina, 2015. 23(4): p. 367–369.

49. Binder, E.B., et al., Association of polymorphisms in genes regulating the corticotropin-releasing factor system with antidepressant treatment response. Archives of general psychiatry, 2010. 67(4): p. 369–379.

50. Murphy, E., et al., Race, genetic ancestry and response to antidepressant treatment for major depression. Neuropsychopharmacology, 2013. 38(13): p. 2598–606.

51. Chaudhry, M., et al., Impact of CYP2D6 genotype on amitriptyline efficacy for the treatment of diabetic peripheral neuropathy: a pilot study. Pharmacogenomics, 2017. 18(5): p. 433–443.

52. Sistonen, J., et al., CYP2D6 worldwide genetic variation shows high frequency of altered activity variants and no continental structure. Pharmacogenet Genomics, 2007. 17(2): p. 93–101.

53. Kirchheiner, J., et al., Effects of the CYP2D6 gene duplication on the pharmacokinetics and pharmacodynamics of tramadol. Journal of Clinical Psychopharmacology, 2008. 28(1): p. 78–83.

54. Bathum, L., et al., Phenotypes and genotypes for CYP2D6 and CYP2C19 in a black Tanzanian population. Br J Clin Pharmacol, 1999. 48(3): p. 395–401.

55. Wright, G.E., et al., Elucidation of CYP2D6 genetic diversity in a unique African population: implications for the future application of pharmacogenetics in the Xhosa population. Ann Hum Genet, 2010. 74(4): p. 340–50.

56. Masimirembwa, C.M., et al., Low CYP1A2 activity in rural Shona children of Zimbabwe. Clin Pharmacol Ther, 1995. 57(1): p. 25–31.

57. Masimirembwa, C.M., et al., Genetic polymorphism of cytochrome P450 CYP2D6 in Zimbabwean population. Pharmacogenetics, 1993. 3(6): p. 275–80.

58. Brosen, K., Some aspects of genetic polymorphism in the biotransformation of antidepressants. Therapie, 2004. 59(1): p. 5–12.

59. Dandara, C., et al., Genetic polymorphism of CYP2D6 and CYP2C19 in East- and Southern African populations including psychiatric patients. European Journal of Clinical Pharmacology, 2001. 57(1): p. 11–17.

60. Persson, I., et al., S-mephenytoin hydroxylation phenotype and CYP2C19 genotype among Ethiopians. Pharmacogenetics, 1996. 6(6): p. 521–6.

61. Sim, S.C., et al., A common novel CYP2C19 gene variant causes ultrarapid drug metabolism relevant for the drug response to proton pump inhibitors and antidepressants. Clin Pharmacol Ther, 2006. 79(1): p. 103–13.

62. Miura, J., et al., Cytochrome P450 2C19 genetic polymorphisms in Ugandans. Eur J Clin Pharmacol, 2009. 65(3): p. 319–20.

63. Masimirembwa, C., et al., Phenotyping and genotyping of S-mephenytoin hydroxylase (cytochrome P450 2C19) in a Shona population of Zimbabwe. Clin Pharmacol Ther, 1995. 57(6): p. 656–61.

64. Nsabiyumva, F., et al., Oxidative polymorphism of dextromethorphan in a Burundi population. Eur J Clin Pharmacol, 1991. 41(1): p. 75–7.

65. Zanger, U.M., et al., Functional pharmacogenetics/genomics of human cytochromes P450 involved in drug biotransformation. Anal Bioanal Chem, 2008. 392(6): p. 1093–108.

66. Mouterde, M., et al., Joint Analysis of Phenotypic and Genomic Diversity Sheds Light on the Evolution of Xenobiotic Metabolism in Humans. Genome Biology and Evolution, 2022. 14(12): p. 23.

67. Motshoge, T., et al., Cytochrome P450 2C8*2 allele in Botswana: Human genetic diversity and public health implications. Acta Trop, 2016. 157: p. 54–8.

68. Bayoumi, R.A.L., et al., The N-acetyltransferase G191 a mutation among Sudanese and Somalis. Pharmacogenetics, 1997. 7(5): p. 397–399.

69. Pavlos, R., S. Mallal, and E. Phillips, HLA and pharmacogenetics of drug hypersensitivity. Pharmacogenomics, 2012. 13(11): p. 1285–306.

70. Ali, A.A., et al., Genome-wide analyses disclose the distinctive HLA architecture and the pharmacogenetic landscape of the Somali population. Sci Rep, 2020. 10(1): p. 5652.

71. Merwe, N.V.D., et al., CYP2D6 genotyping and use of antidepressants in breast cancer patients: Test development for clinical application. Metabolic Brain Disease, 2012. 27(3): p. 319–326.

72. Dodgen, T.M., et al., Risperidone-associated adverse drug reactions and CYP2D6 polymorphisms in a South African cohort. Applied and Translational Genomics, 2015. 5: p. 40–46.

73. Ajmi, M., et al., Association between ABCB1 polymorphisms and response to first-generation antiepileptic drugs in a Tunisian epileptic population. Int J Neurosci, 2018. 128(8): p. 705–714.

74. Santovito, A., et al., Polymorphisms of cytochrome P450 1A1, glutathione s-transferases M1 and T1 genes in Ouangolodougou (Northern Ivory Coast). Genetics and Molecular Biology, 2010. 33(3): p. 434–437.

75. Gyamfi, M.A., et al., High prevalence of cytochrome P450 2A6*1A alleles in a black African population of Ghana. Eur J Clin Pharmacol, 2005. 60(12): p. 855–7.

76. Xie, H.G., et al., Genetic polymorphism of (S)-mephenytoin 4’-hydroxylation in populations of African descent. British Journal of Clinical Pharmacology, 1999. 48(3): p. 402–408.

77. Griese, E.U., et al., Analysis of the CYP2D6 gene mutations and their consequences for enzyme function in a West African population. Pharmacogenetics, 1999. 9(6): p. 715–23.

78. Horsmans, Y., J.M. Kanyinda, and J.P. Desager, Relationship between mephenytoin, phenytoin and tolbutamide hydroxylations in healthy African subjects. Pharmacology and Toxicology, 1996. 78(2): p. 86–88.

79. Afilal, D., et al., Genetic Polymorphism of Drug-Metabolizing Enzymes CYP2C9 and CYP2C19 in Moroccan Population. Genet Test Mol Biomarkers, 2017. 21(5): p. 298–304.

80. Adehin, A., et al., Relationship between metabolic phenotypes and genotypes of CYP1A2 and CYP2A6 in the Nigerian population. Drug Metab Pers Ther, 2017. 32(1): p. 39–47.

81. Adehin, A., O.O. Bolaji, and M.A. Kennedy, Polymorphisms in CYP2C8 and CYP3A5 genes in the Nigerian population. Drug Metab Pharmacokinet, 2017. 32(3): p. 189–191.

82. Gomez, F., J. Hirbo, and S.A. Tishkoff, Genetic variation and adaptation in Africa: implications for human evolution and disease. Cold Spring Harb Perspect Biol, 2014. 6(7): p. a008524.

83. Mathuba, B., et al., Catalyzing clinical implementation of pharmacogenomics and personalized medicine interventions in Africa. Pharmacogenomics, 2021. 22(2): p. 115–122.

84. Tiwari, A.K., et al., A common polymorphism in the cannabinoid receptor 1 (CNR1) gene is associated with antipsychotic-induced weight gain in Schizophrenia. Neuropsychopharmacology, 2010. 35(6): p. 1315–24.

85. Ravyn, D., et al., CYP450 pharmacogenetic treatment strategies for antipsychotics: a review of the evidence. Schizophr Res, 2013. 149(1-3): p. 1–14.

86. Cacabelos, R. and R. Martinez-Bouza, Genomics and pharmacogenomics of schizophrenia. CNS Neurosci Ther, 2011. 17(5): p. 541–65.

87. Coleman, J.J. and S.K. Pontefract, Adverse drug reactions. Clin Med (Lond), 2016. 16(5): p. 481–485.

88. Hendset, M., et al., The complexity of active metabolites in therapeutic drug monitoring of psychotropic drugs. Pharmacopsychiatry, 2006. 39(4): p. 121–7.

89. Bochud, M. and I. Guessous, Gene-environment interactions of selected pharmacogenes in arterial hypertension. Expert Rev Clin Pharmacol, 2012. 5(6): p. 677–86.

90. Mpye, K.L., et al., Disease burden and the role of pharmacogenomics in African populations. Glob Health Epidemiol Genom, 2017. 2: p. e1.

91. Greden, J.F., et al., Impact of pharmacogenomics on clinical outcomes in major depressive disorder in the GUIDED trial: A large, patient- and rater-blinded, randomized, controlled study. J Psychiatr Res, 2019. 111: p. 59–67.

92. Kaniwa, N., et al., HLA-B*1511 is a risk factor for carbamazepine-induced Stevens-Johnson syndrome and toxic epidermal necrolysis in Japanese patients. Epilepsia, 2010. 51(12): p. 2461–5.

93. Hall-Flavin, D.K., et al., Utility of integrated pharmacogenomic testing to support the treatment of major depressive disorder in a psychiatric outpatient setting. Pharmacogenet Genomics, 2013. 23(10): p. 535–48.

94. Jukic, M.M., et al., Impact of CYP2C19 Genotype on Escitalopram Exposure and Therapeutic Failure: A Retrospective Study Based on 2,087 Patients. Am J Psychiatry, 2018. 175(5): p. 463–470.

95. da Rocha, J.E.B., et al., The Extent and Impact of Variation in ADME Genes in Sub-Saharan African Populations. Front Pharmacol, 2021. 12: p. 634016.

96. Mukonzo, J.K., The challenge of paediatric efavirenz dosing: implications and way forward for the sub-Saharan Africa. AIDS, 2014. 28(13): p. 1855–7.

97. Dandara, C., et al., Cytochrome P450 pharmacogenetics in African populations: implications for public health. Expert Opin Drug Metab Toxicol, 2014. 10(6): p. 769–85.

98. Awandu, S.S., et al., Understanding human genetic factors influencing primaquine safety and efficacy to guide primaquine roll-out in a pre-elimination setting in southern Africa. Malar J, 2018. 17(1): p. 120.

99. Goulding, R., et al., Genotype-guided drug prescribing: a systematic review and meta-analysis of randomized control trials. Br J Clin Pharmacol, 2015. 80(4): p. 868–77.

100. Shad, M.U., Genetic Testing for Antipsychotic Pharmacotherapy: Bench to Bedside. Behav Sci (Basel), 2021. 11(7).

101. Franco-Martin, M.A., et al., Usefulness of Pharmacogenetic Analysis in Psychiatric Clinical Practice: A Case Report. Clin Psychopharmacol Neurosci, 2018. 16(3): p. 349–357.

102. Dodgen, T.M., et al., Risperidone-associated adverse drug reactions and CYP2D6 polymorphisms in a South African cohort. Appl Transl Genom, 2015. 5: p. 40–6.

103. Marigorta, U.M., et al., Replicability and Prediction: Lessons and Challenges from GWAS. Trends Genet, 2018. 34(7): p. 504–517.

104. Ertekin-Taner, N., Genetics of Alzheimer disease in the pre- and post-GWAS era. Alzheimers Res Ther, 2010. 2(1): p. 3.

105. Adiukwu, F., et al., Psychiatry pharmacogenomics: Africans are not at the table. Lancet Psychiatry, 2023. 10(2): p. 80.

106. Florence Femi Odekunle, R.O.O., Srinivasan Shankar, Why sub-Saharan Africa lags in electronic health record adoption and possible strategies to increase its adoption in this region. International Journal of Health Sciences-Ijhs, 2017. 11(4).

107. Zhong, A., et al., Ethical, social, and cultural issues related to clinical genetic testing and counseling in low- and middle-income countries: a systematic review. Genet Med, 2021. 23(12): p. 2270–2280.

108. Karen Maigetter, A.M.P., Abhay Kadam, Kim Ward, Mitchell G. Weiss, Pharmacovigilance in India, Uganda and South Africa with reference to WHO’s minimum requirements. International Journal of Health Policy and Management, 2015. 4(5): p. 295–305.

109. June C. Carroll, A.L.R., Brenda J. Wilson, Judith Allanson, Sean M. Blaine, Mary Jane Esplen, Sandra A. Farrell, Gail E. Graham, Jennifer MacKenzie, Wendy Meschino, Fiona Miller, Preeti Prakash, Cheryl Shuman, Anne Summers, Sherry Taylor, Genetic education for primary care providers:Improving attitudes, knowledge, and confidence. Canadian Family Physician, 2009. 55.

110. Ogunrin, O., F. Taiwo, and L. Frith, Genomic Literacy and Awareness of Ethical Guidance for Genomic Research in Sub-Saharan Africa: How Prepared Are Biomedical Researchers? J Empir Res Hum Res Ethics, 2019. 14(1): p. 78–87.

111. Moldrup, C., Ethical, social and legal implications of pharmacogenomics: a critical review. Community Genet, 2001. 4(4): p. 204–14.

112. Mahomed, S., Human Biobanking in Developed and Developing Countries: An Ethico-Legal Comparative Analysis of the Frameworks in the United Kingdom, Australia, Uganda, and South Africa. Camb Q Healthc Ethics, 2021. 30(1): p. 146–160.

113. Veenstra, D. and W. Burke, Pharmacogenomics and public health. Public Health Genomics, 2009. 12(3): p. 131–3.

114. Rodriguez-Laso, A., et al., A Map of the Initiatives That Harmonize Patient Cohorts Across the World. Front Public Health, 2021. 9: p. 666844.

115. Gomez-Olive, F.X., et al., Cohort Profile: Health and Ageing in Africa: A Longitudinal Study of an INDEPTH Community in South Africa (HAALSI). Int J Epidemiol, 2018. 47(3): p. 689–690j.

116. Suissa, S., E.E.M. Moodie, and S. Dell’Aniello, Prevalent new-user cohort designs for comparative drug effect studies by time-conditional propensity scores. Pharmacoepidemiology and Drug Safety, 2017. 26(4): p. 459–468.

117. Mekonnen, A.B., et al., Adverse Drug Events and Medication Errors in African Hospitals: A Systematic Review. Drugs - Real World Outcomes, 2017. 5(1): p. 1–24.

118. Dandara, C., et al., African Pharmacogenomics Consortium: Consolidating pharmacogenomics knowledge, capacity development and translation in Africa: Consolidating pharmacogenomics knowledge, capacity development and translation in Africa. AAS Open Res, 2019. 2: p. 19.

119. Genetics, A.S.o.H., Human Heredity & Health in Africa (H3Africa). 2020: South Africa.

120. Mboowa, G., et al., The Collaborative African Genomics Network (CAfGEN): Applying Genomic technologies to probe host factors important to the progression of HIV and HIV-tuberculosis infection in sub-Saharan Africa. AAS Open Res, 2018. 1: p. 3.

121. Obamba, M.O. and J.K. Mwema, Symmetry and Asymmetry: New Contours, Paradigms, and Politics in African Academic Partnerships. Higher Education Policy, 2009. 22(3): p. 349–371.

122. Munung, N.S., B.M. Mayosi, and J. de Vries, Equity in international health research collaborations in Africa: Perceptions and expectations of African researchers. PLoS One, 2017. 12(10): p. e0186237.

123. Chowdhury, A. and M. Fakruddin, Pharmacogenomics-The Promise of Personalized Medicine. Bangladesh Journal of Medical Science, 2013. 12(4): p. 346–356.

124. Zycher, B., J.A. DiMasi, and C.P. Milne, Private sector contributions to pharmaceutical science: thirty-five summary case histories. Am J Ther, 2010. 17(1): p. 101–20.

125. Sharawy, I., Ethical and Regulatory Issues in Genetic Research Collaboration between Global Pharmaceutical Industry and Low- and Middle-income Countries. Research Square, 2022.

